# Biomechanical characterization of head and neck tumors and corresponding lymph nodes for nodal assessment by ultrasound shear wave elastography and MR elastography

**DOI:** 10.64898/2026.11.12.26367958

**Authors:** Pedro Augusto Dantas de Moraes, Mehrgan Shahryari, Steffen Dommerich, Heidi Olze, Jonathan Pohl, Iris Piwonski, Thomas Fischer, Weijie Zhao, Kudor Harb, Jing Guo, Ingolf Sack, Katharina Margherita Wakonig

## Abstract

**Background:** Malignant remodeling alters tissue mechanics, but biomechanical differences among primary head and neck tumors, benign cervical lymph nodes, and nodal metastases remain poorly characterized. We used in vivo and ex vivo ultrasound shear-wave elastography (USE) and ex vivo tabletop MR elastography (ttMRE) to compare stiffness, viscoelasticity, and spatial heterogeneity across these tissue groups and to assess cross-modal agreement. As a secondary objective, we explored whether primary-tumor biomechanics differed by pathological nodal status.

**Methods:** Eighteen patients - 17 with head and neck squamous cell carcinoma (HNSCC) and one with salivary duct carcinoma - underwent surgical resection. Target cervical lymph nodes were assessed by in vivo USE, and resected lymph nodes and primary tumors by ex vivo USE. Multifrequency ttMRE was successfully performed in 8 benign lymph nodes, 8 metastatic lymph nodes, and 13 primary tumors and yielded shear modulus μ, power-law exponent α, and spatial heterogeneity metrics. Quantitative T1, T2, fat-fraction, and apparent-diffusion-coefficient (ADC) measurements were also performed. Group comparisons used Kruskal-Wallis tests with Dunn-Bonferroni post-hoc tests, paired comparisons used Wilcoxon signed-rank tests, and primary tumors from node-positive versus node-negative patients were compared using Mann-Whitney U tests. Diagnostic performance was summarized by area under the receiver operating characteristic curve (AUC), and cross-modal agreement by Spearman correlation.

**Results:** By ttMRE, benign lymph nodes were softer than metastatic lymph nodes and primary tumors (median μ [IQR], 3.4 [3.1-8.9] vs 57.3 [26.2-102.4] vs 24.2 [13.2-37.6] kPa; p = 0.001) with more homogeneous texture as indicated by lower stiffness entropy (H(μ), 1.24 [1.05-1.30] vs 1.88 [1.71-2.27] vs 1.70 [1.56-1.78] nats; p = 0.002). Although the unpaired distributions of metastatic nodes and primary tumors overlapped, matched within-patient comparisons showed higher stiffness in primary tumors than in benign nodes and in metastatic nodes than in their corresponding primary tumors (n = 6 pairs per comparison; p = 0.031 for each). For discriminating metastatic from benign lymph nodes, AUC was 0.97 for ttMRE μ, 0.96 for in vivo USE, and 0.90 for ex vivo USE. By ttMRE, primary tumors from node-positive patients had lower stiffness than those from node-negative patients (12.1 [10.2-16.1] vs 30.1 [26.4-72.8] kPa; uncorrected p = 0.035), whereas ex vivo USE did not show differences (p = 0.415). ttMRE μ correlated with ex vivo USE (ρ = 0.70, p < 0.001) and in vivo (ρ = 0.69, p = 0.003).

**Conclusion:** In our prospective cohort, tissue stiffness was the main biomechanical feature distinguishing benign from metastatic cervical lymph nodes. Furthermore, an association between lower primary-tumor stiffness and pathological nodal positivity was observed by MRE but not USE. Our findings establish a proof of concept for the biomechanical characterization of primary head and neck tumors and regional lymph nodes.

## 1. Introduction

Tumor biomechanics emerge from interactions among malignant cells, extracellular matrix, vasculature, and interstitial fluid. Matrix deposition, cross-linking, and fiber alignment can increase bulk tissue stiffness, while malignant cells within the same tumor may remain mechanically soft and motile; the measured phenotype therefore depends on spatial scale, loading, and imaging method [1–4]. Viscoelasticity and spatial heterogeneity may capture additional aspects of this remodeling that are not represented by anatomy alone [5, 6].

In head and neck malignancies, cervical nodal status is central to staging and treatment planning. Although cross-sectional imaging with CT, MRI, or FDG-PET/CT is mandatory to assess the size and local extent of the primary tumor as well as the presence of distant metastases, additional cervical ultrasonography is recommended for the assessment of cervical lymph nodes because of its higher diagnostic performance [7–9]. Nevertheless, nodal evaluation continues to rely substantially on size and morphological criteria. USE adds a quantitative mechanical readout. Pathology-referenced studies have generally reported higher stiffness in metastatic than in benign cervical nodes, although thresholds vary with the elastography technique, region-of-interest strategy, and nodal composition [10–12]. USE is clinically accessible but remains sensitive to operator, geometry, depth, and boundary conditions and usually reduces complex tissue behavior to an effective shear-wave speed or stiffness value [13, 14].

MRE quantifies tissue mechanics by imaging externally induced shear-wave propagation [15–17]. Across several tumor entities, MRE-derived parameters have been associated with lesion type, grade, treatment response, recurrence, and other adverse pathological features [18–29]. Multifrequency MRE also characterizes wave dispersion and attenuation, providing stiffness- and fluidity-related parameters that depend on tissue microstructure [6, 23–26].

Head-and-neck MRE studies have established feasibility in the parotid glands and cervical region [30–35]. Multiparametric ultrasound incorporating shear-wave elastography has also shown potential for preoperative parotid-tumor assessment [36]. Higher stiffness has been reported in metastatic lymph nodes using in vivo multifrequency MRE in the abdomen and DWI-based ‘virtual stiffness’ in the neck [37, 38]. However, MRE has not been used to characterize resected cervical nodes together with matched primary tumors while benchmarking the same specimens against in vivo and ex vivo USE.

Tabletop MRE (ttMRE) enables high-frequency, high-resolution measurements in small excised specimens under controlled experimental conditions [39–42]. Prior ex vivo work has shown sensitivity to tissue composition, tumor-cell viability, treatment response, and matrix-cell interactions [35, 43–46]. ttMRE therefore provides a research platform for detailed mechanical phenotyping, but its measurements are not directly interchangeable with in vivo elastography because frequency range and boundary conditions differ.

In this prospective study, we combined in vivo and ex vivo USE with ex vivo ttMRE and quantitative T1, T2, fat-fraction, and ADC mapping. The primary objectives were to compare biomechanical parameters among benign cervical lymph nodes, metastatic cervical lymph nodes, and primary tumors and to assess agreement between ttMRE stiffness and USE shear-wave speed. Secondary, exploratory objectives were to evaluate whether primary-tumor biomechanics differed by pathological nodal status and to describe associations with quantitative MRI and qualitative histopathology.

## 2. Materials and Methods

### 2.1 Study cohort and specimen inclusion

Eighteen patients with histopathologically confirmed head and neck malignancies who were scheduled for surgical resection were prospectively enrolled (Fig. 1). One day before surgery, the operating surgeon performed in vivo USE of the target cervical lymph node and subsequently identified and retrieved the same node during neck dissection. According to the study protocol, all resected target lymph nodes and primary-tumor specimens were scheduled for immediate ex vivo USE and ttMRE; when tissue volume was limited, diagnostic histopathology took priority. Final histopathology confirmed head and neck squamous cell carcinoma (HNSCC) in 17 patients and salivary duct carcinoma in one.

**Figure 1.**
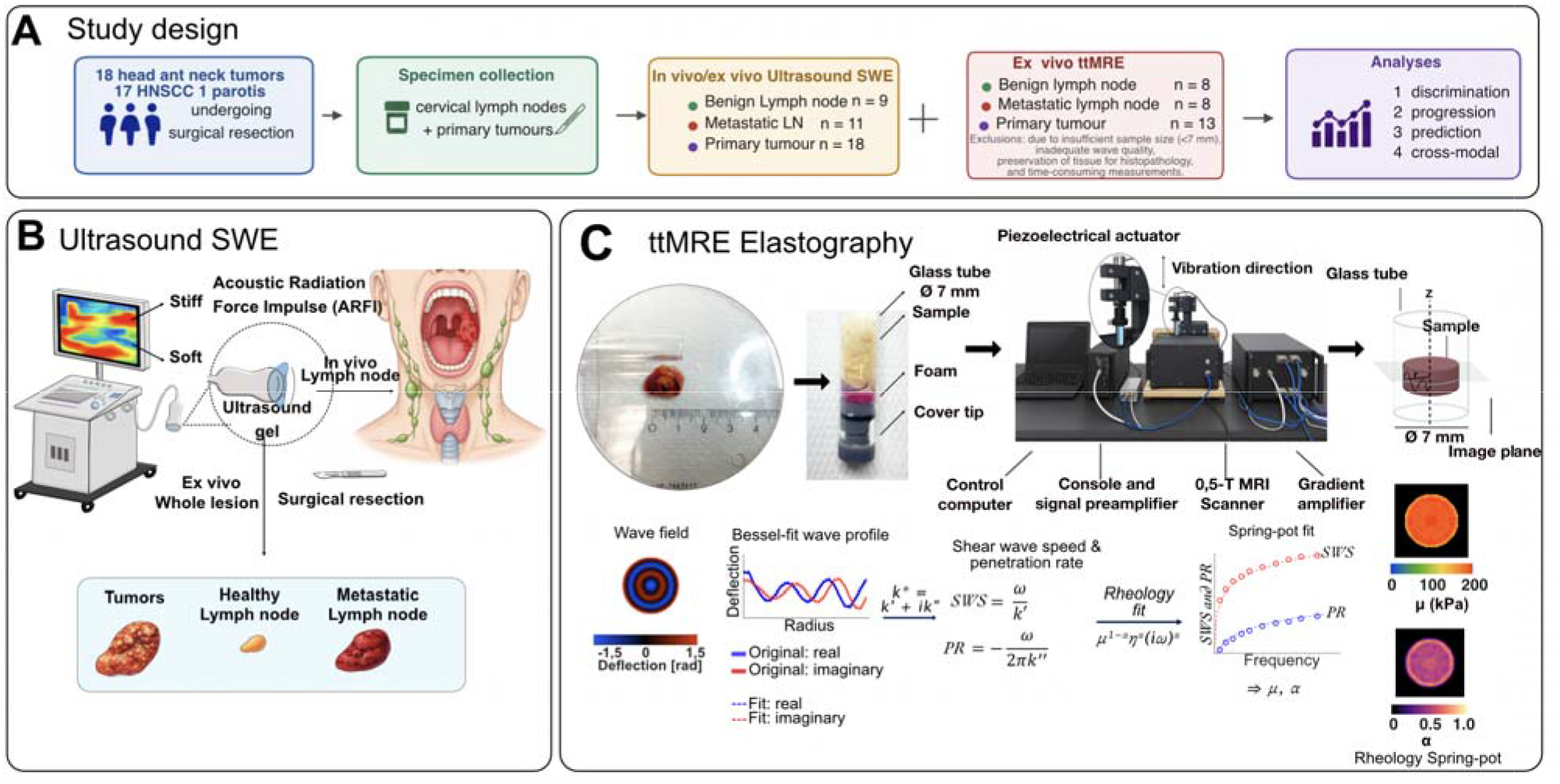
Study design and elastography methods. Eighteen enrolled patients (17 HNSCC and one salivary duct carcinoma) underwent surgical resection. (A) Workflow and sequence-specific analysis cohorts. USE was evaluable in 9 benign lymph nodes, 11 metastatic lymph nodes, and 18 primary tumors; ttMRE was evaluable after exclusions in 8 benign lymph nodes, 8 metastatic lymph nodes, and 13 primary tumors (7 node-negative and 6 node-positive). (B) ARFI-based USE: target cervical lymph nodes were examined in vivo before resection, and resected nodes and primary tumors were examined ex vivo immediately after surgery. (C) ttMRE: specimens were placed in a 7-mm glass tube, vibrated on a 0.5-T tabletop MRI system, and analyzed using Bessel-informed reconstruction of frequency-resolved shear-wave speed (SWS, m/s) and penetration rate (PR, m/s) followed by pixel-wise spring-pot fitting to obtain shear modulus μ and power-law exponent α. Exclusion reasons are detailed in Section 3.1.

The study was approved by the institutional ethics committee of Charité-Universitätsmedizin Berlin (protocol code EA4/214/23; date of approval: November 27, 2024), and written informed consent was obtained from all participants according to the Declaration of Helsinki.

### 2.2 Ultrasound shear-wave elastography

All ultrasound examinations followed a standardized protocol using an Aplio i900 system (Canon Medical Systems Corporation, Tochigi, Japan) with a 4-10 MHz multifrequency linear-array transducer (center frequency, 7 MHz) [13, 14]. Patients were examined supine with the head extended and rotated contralaterally. B-mode ultrasound was used to document target-node location, morphology, and short- and long-axis diameters.

USE based on acoustic radiation force impulse imaging was then performed in vivo. Localized acoustic push pulses generated shear waves within the target node, and three consecutive two-dimensional acquisitions were obtained with the node centered in the field of view. A circular region of interest (ROI) was manually placed within the node, and its size was adapted to the lesion. Shear-wave speed (SWS, m/s) maps were generated, and the mean SWS and corresponding standard deviation across the three acquisitions were recorded. Dual-image display of the B-mode image and color-coded USE map was used to verify lesion coverage. Penetration depth was adapted to the individual node, whereas gain settings were held constant. SWS was reported directly in m/s rather than converted to Young’s modulus. The commonly used conversion is

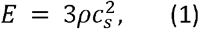

where E is Young’s modulus, ρ is tissue density, and c□ is the SWS. Equation (1) assumes homogeneous, isotropic, incompressible, and purely elastic tissue. These assumptions are not generally valid in structurally heterogeneous cervical lymph nodes; direct reporting of SWS therefore avoids model-dependent inaccuracies [13, 14].

All in vivo examinations were performed one day before surgery by the surgeon who subsequently performed the tumor resection and neck dissection, facilitating reliable intraoperative identification of the target node. The examiner held German Society for Ultrasound in Medicine (DEGUM) Level II certification, corresponding approximately to European Federation of Societies for Ultrasound in Medicine and Biology (EFSUMB) Level 2 competency in head and neck ultrasound [47].

Immediately after resection, the target lymph node and primary-tumor specimens underwent ex vivo USE using the same ARFI-based protocol. Each resected specimen was examined once. Mean SWS within a manually defined ROI was recorded as the ex vivo USE stiffness readout.

### 2.3 Tabletop MR elastography: setup and acquisition

ttMRE was performed on a benchtop 0.5-T MRI system (Magspec, Pure Devices GmbH, Würzburg, Germany) equipped with a four-channel gradient amplifier (DC-600, Pure Devices GmbH) and a piezoelectric actuator (PAHL60/20, Piezosystem Jena GmbH). The actuator generated vibrations along the longitudinal tube axis (Fig. 1C). Fresh specimens were placed in cylindrical glass tubes with a 7-mm inner diameter and mechanically coupled using a PVC-foam spacer. A spin-echo sequence with bipolar motion-encoding gradients directed along z encoded one 3-mm axial slice at 0.15 × 0.15 mm² in-plane resolution (64 × 64 matrix) at four phase offsets per vibration period. The actuator was driven from 800 to 5000 Hz; 800 Hz yielded no evaluable wave field in any specimen, so retained frequencies spanned 900 to 5000 Hz, and in most specimens the retained range was 900-3300 Hz (Supplementary Table S1). Echo time, repetition time, and motion-encoding-gradient amplitude were adjusted specimen-wise according to relaxation behavior. This subsection describes acquisition only; processing is detailed below. The hardware and acquisition principles follow Zhao et al. [41].

### 2.4 ttMRE post-processing and spring-pot rheological modeling

The complex wave field at each drive frequency was obtained after phase unwrapping and temporal Fourier transformation. First, a global Bessel-function fit to the complex radial wave profile provided a specimen-level complex wavenumber and a prior for spatial filtering (Fig. 3). Directional bandpass filtering then isolated quasi-plane shear-wave components along eight directions. From the spatial phase and amplitude gradients of the filtered complex fields, frequency-resolved maps of SWS(f) and penetration rate PR(f), both in m/s, were reconstructed pixel-wise [41, 48–50]. The paired use of SWS and PR across frequencies follows wideband viscoelastic characterization principles also used in time-harmonic ultrasound elastography [51]. In contrast to frequency- and amplitude-weighted averaged maps described for other applications of this framework, no frequency-averaged SWS or PR maps were used here. Instead, the frequency-resolved SWS(f) and PR(f) values at each pixel were fitted directly with a fractional spring-pot model to obtain shear modulus μ (reported in kPa; hereafter also referred to as stiffness) and power-law exponent α (dimensionless; α → 0 indicates more elastic and α → 1 more viscous behavior). The spring-pot model was defined as

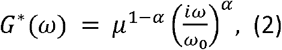

where G* is the complex shear modulus, ω is angular frequency (ω = 2πf), ω₀ is the reference angular frequency, μ is the shear modulus at the reference frequency f₀ = 1000 Hz (ω₀ = 2πf₀), and 0 ≤ α ≤ 1 describes the transition from elastic-like (α → 0) to viscous-like (α → 1) behavior.

**Table 1.**
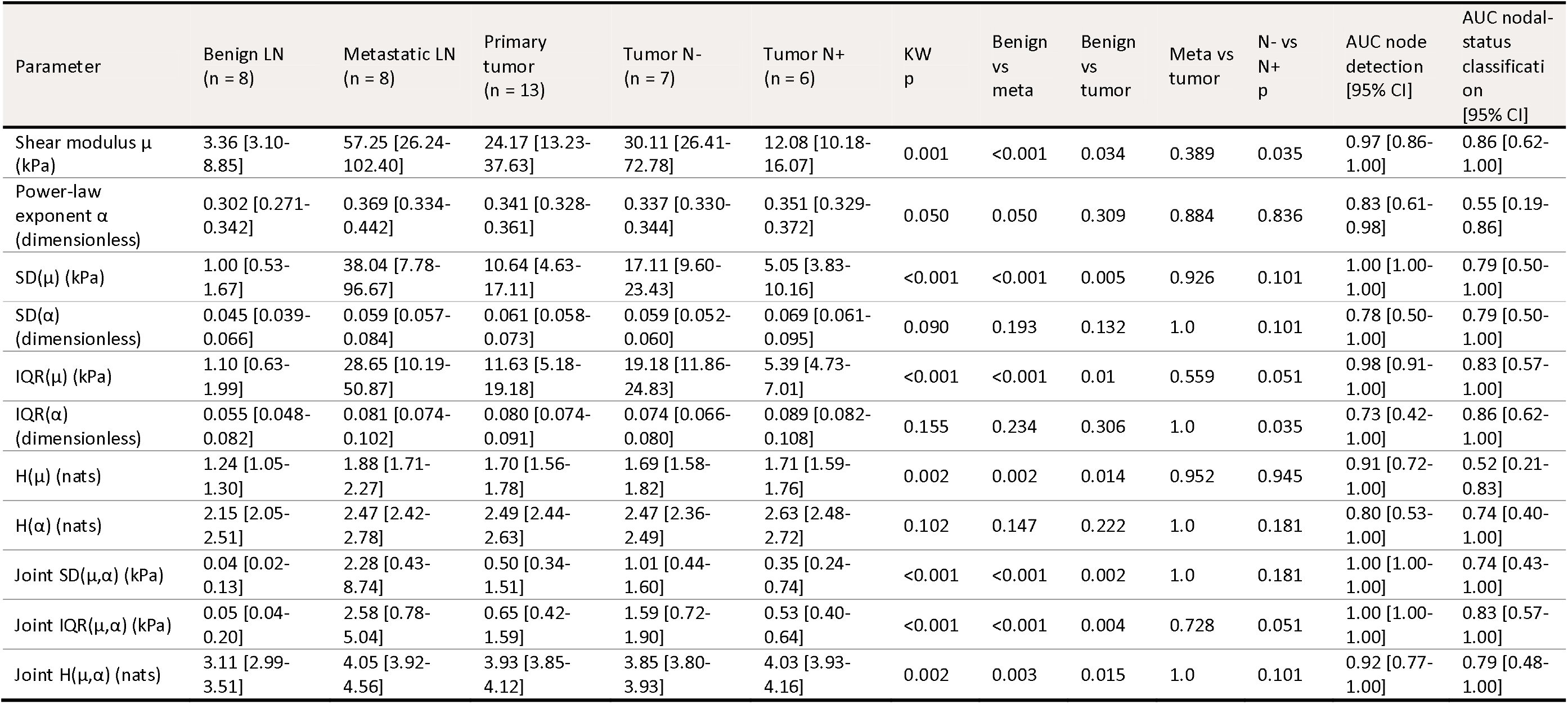
Tabletop MR elastography. Biomechanical parameters and heterogeneity metrics. The table reports shear modulus μ, power-law exponent α, and heterogeneity metrics for benign nodes, metastatic nodes, and primary tumors, with primary tumors stratified by pathological nodal status. H is Shannon entropy (natural logarithm; nats); joint IQR and joint SD are products of the μ and α terms. Values are medians [IQR]; AUC values include 2000-resample bootstrap 95% confidence intervals. p values below 0.05 are bold. Abbreviations: LN, lymph node; N-, node-negative; N+, node-positive; SD, standard deviation; IQR, interquartile range; AUC, area under the ROC curve; CI, confidence interval.

| Parameter | Benign LN<br>(n = 8) | Metastatic LN<br>(n = 8) | Primary tumor<br>(n = 13) | Tumor N-<br>(n = 7) | Tumor N+<br>(n = 6) | KW<br>p | Benign vs meta | Benign vs tumor | Meta vs tumor | N- vs N+<br>p | AUC node detection<br>[95% CI] | AUC nodal-status classification<br>[95% CI] |
|---|---|---|---|---|---|---|---|---|---|---|---|---|
| Shear modulus μ (kPa) | 3.36 [3.10-8.85] | 57.25 [26.24-102.40] | 24.17 [13.23-37.63] | 30.11 [26.41-72.78] | 12.08 [10.18-16.07] | 0.001 | <0.001 | 0.034 | 0.389 | 0.035 | 0.97 [0.86-1.00] | 0.86 [0.62-1.00] |
| Power-law exponent α (dimensionless) | 0.302 [0.271-0.342] | 0.369 [0.334-0.442] | 0.341 [0.328-0.361] | 0.337 [0.330-0.344] | 0.351 [0.329-0.372] | 0.050 | 0.050 | 0.309 | 0.884 | 0.836 | 0.83 [0.61-0.98] | 0.55 [0.19-0.86] |
| SD(μ) (kPa) | 1.00 [0.53-1.67] | 38.04 [7.78-96.67] | 10.64 [4.63-17.11] | 17.11 [9.60-23.43] | 5.05 [3.83-10.16] | <0.001 | <0.001 | 0.005 | 0.926 | 0.101 | 1.00 [1.00-1.00] | 0.79 [0.50-1.00] |
| SD(α) (dimensionless) | 0.045 [0.039-0.066] | 0.059 [0.057-0.084] | 0.061 [0.058-0.073] | 0.059 [0.052-0.060] | 0.069 [0.061-0.095] | 0.090 | 0.193 | 0.132 | 1.0 | 0.101 | 0.78 [0.50-1.00] | 0.79 [0.50-1.00] |
| IQR(μ) (kPa) | 1.10 [0.63-1.99] | 28.65 [10.19-50.87] | 11.63 [5.18-19.18] | 19.18 [11.86-24.83] | 5.39 [4.73-7.01] | <0.001 | <0.001 | 0.01 | 0.559 | 0.051 | 0.98 [0.91-1.00] | 0.83 [0.57-1.00] |
| IQR(α) (dimensionless) | 0.055 [0.048-0.082] | 0.081 [0.074-0.102] | 0.080 [0.074-0.091] | 0.074 [0.066-0.080] | 0.089 [0.082-0.108] | 0.155 | 0.234 | 0.306 | 1.0 | 0.035 | 0.73 [0.42-1.00] | 0.86 [0.62-1.00] |
| H(μ) (nats) | 1.24 [1.05-1.30] | 1.88 [1.71-2.27] | 1.70 [1.56-1.78] | 1.69 [1.58-1.82] | 1.71 [1.59-1.76] | 0.002 | 0.002 | 0.014 | 0.952 | 0.945 | 0.91 [0.72-1.00] | 0.52 [0.21-0.83] |
| H(α) (nats) | 2.15 [2.05-2.51] | 2.47 [2.42-2.78] | 2.49 [2.44-2.63] | 2.47 [2.36-2.49] | 2.63 [2.48-2.72] | 0.102 | 0.147 | 0.222 | 1.0 | 0.181 | 0.80 [0.53-1.00] | 0.74 [0.40-1.00] |
| Joint SD(μ,α) (kPa) | 0.04 [0.02-0.13] | 2.28 [0.43-8.74] | 0.50 [0.34-1.51] | 1.01 [0.44-1.60] | 0.35 [0.24-0.74] | <0.001 | <0.001 | 0.002 | 1.0 | 0.181 | 1.00 [1.00-1.00] | 0.74 [0.43-1.00] |
| Joint IQR(μ,α) (kPa) | 0.05 [0.04-0.20] | 2.58 [0.78-5.04] | 0.65 [0.42-1.59] | 1.59 [0.72-1.90] | 0.53 [0.40-0.64] | <0.001 | <0.001 | 0.004 | 0.728 | 0.051 | 1.00 [1.00-1.00] | 0.83 [0.57-1.00] |
| Joint H(μ,α) (nats) | 3.11 [2.99-3.51] | 4.05 [3.92-4.56] | 3.93 [3.85-4.12] | 3.85 [3.80-3.93] | 4.03 [3.93-4.16] | 0.002 | 0.003 | 0.015 | 1.0 | 0.101 | 0.92 [0.77-1.00] | 0.79 [0.48-1.00] |

Quantitative analysis was restricted to the central 5.6-mm region of the 7-mm tube to reduce boundary effects. Frequencies were retained only when the central analysis region showed adequate wave amplitude and phase signal-to-noise ratio and the wavelength was sufficiently sampled; frequencies were excluded when attenuation reduced the wave amplitude to the noise floor before the wave reached the analysis region or when the wavelength could not be resolved reliably [41]. Spatially isolated fitting failures in μ and α were corrected using an iterative log-space local-median filter. A pixel was classified as a fitting failure when, after log10 transformation of the parameter map, it deviated from the median of its 5 × 5 neighborhood by more than four robust standard deviations (1.4826 × median absolute deviation, with a lower bound of 0.05 log10 units) and simultaneously differed from that local median by more than a factor of 20; pixels deviating from the specimen-wide median by more than five robust standard deviations and meeting the same factor-20 criterion were also flagged. At least eight valid neighboring pixels were required for a pixel to be evaluated. Flagged pixels were replaced by the geometric median of the surrounding non-flagged pixels, with the window expanded from 5 × 5 to at most 21 × 21 until this minimum was reached, and the procedure was repeated for up to four iterations or until no further pixels were flagged. Because a statistical and a ratio criterion had to be met simultaneously, only isolated implausible values were replaced and spatially coherent heterogeneity was preserved; the proportion of corrected pixels was recorded for each specimen.

### 2.5 Spatial heterogeneity and joint metrics

Spatial heterogeneity (texture) was quantified for μ and α using the within-specimen standard deviation (SD), interquartile range (IQR), and Shannon entropy [5, 41]. Shannon entropy was calculated as

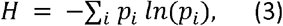

where *p_i_* is the fraction of pixels in histogram bin *i* (number of pixels in bin *i* divided by the total number of analyzed pixels) and ln denotes the natural logarithm; H is therefore reported in nats. The notations H(μ) and H(α) denote univariate entropy, and H(μ,α) denotes joint entropy from the two-dimensional histogram of paired μ-α pixels. Thirty bins were used for each univariate histogram. Because μ spanned several orders of magnitude, its bin edges were defined in log10-space from the pooled 0.5th-99.5th percentiles; using log10 for bin placement does not change the natural-logarithm base in Equation (3). α was binned linearly. Joint IQR and joint SD were defined as the products of the corresponding μ and α metrics, while joint Shannon entropy was calculated from a 30 × 30 two-dimensional histogram using the same log-scaled μ axis.

### 2.6 Multiparametric MRI

A partially overlapping set of resected specimens underwent quantitative ex vivo MRI on the same 0.5-T tabletop system (Fig. 4, Table 3). T1, T2, and fat fraction were evaluable in the same 33 specimens (8 benign lymph nodes, 10 metastatic lymph nodes, 15 primary tumors); ADC was evaluable in 33 specimens (8 benign lymph nodes, 10 metastatic lymph nodes, 15 primary tumors). Every multiparametric MRI cohort was contained within the corresponding USE cohort (9 benign lymph nodes, 11 metastatic lymph nodes, 18 primary tumors). Thus, these data were not a strict subset of the final ttMRE analysis cohort because quantitative MRI remained technically evaluable in some specimens excluded from rheological mapping. T1 relaxation time was measured using inversion-recovery spin-echo imaging with monoexponential fitting, T2 relaxation time using multi-echo spin-echo imaging, proton-density fat fraction using chemical-shift-encoded Dixon-type imaging, and ADC using diffusion-weighted imaging (b values of 0, 27.8, 111.1, 250, 444.4, 694.4, and 1000 s/mm²) with monoexponential signal fitting. Acquisition and processing followed the multiparametric ex vivo tabletop-MRI framework described by Domka et al. [35]. These MRI parameters are reported separately from the ttMRE-derived μ and α.

### 2.7 Histopathological analysis

Formalin-fixed, paraffin-embedded sections from the same specimens were stained with hematoxylin and eosin (H&E), Alcian blue, and Sirius red, and immunostained for matrix metalloproteinase-9 (MMP-9). Specimen processing and whole-slide digitalization were performed at the Institute of Pathology, Charité - Universitätsmedizin Berlin. Whole-slide images were reviewed in QuPath v0.4.3 and assessed qualitatively by visual inspection; no image-based measurements were extracted and no numerical histological values were derived. H&E sections were inspected for tissue architecture and for the apparent density and spatial uniformity of the cellular infiltrate. Alcian blue was used to visualize glycosaminoglycan (GAG) accumulation and Sirius red to visualize collagen deposition and the apparent thickness of collagen fiber bundles. For illustrative purposes, color-coded fiber-orientation overlays of the Sirius red sections were generated with the OrientationJ plugin for ImageJ; these overlays served to display the predominant fiber direction and the visual degree of fiber alignment and were not used to compute orientation statistics. MMP-9 immunostaining was inspected as a marker associated with extracellular-matrix degradation and remodeling. Histological observations were evaluated descriptively.

### 2.8 Statistical analysis

Analyses were conducted at the specimen level except for matched comparisons. Continuous data are reported as median [IQR]. Three-group comparisons used Kruskal-Wallis tests followed, when significant, by Dunn tests with Bonferroni correction for three pairwise contrasts. In vivo USE and primary-tumor comparisons by nodal status used two-sided Mann-Whitney U tests; matched specimens used two-sided Wilcoxon signed-rank tests. Receiver operating characteristic (ROC) curves summarized metastatic-versus-benign node discrimination and, secondarily, nodal-status discrimination from primary tumors. Area under the curve (AUC) confidence intervals were estimated by 2000-resample stratified bootstrap, and Youden thresholds were calculated in the study cohort. AUCs were not formally compared. Cross-modal associations were analyzed by Spearman correlation. To account for repeated specimens from individual patients, tissue-class comparisons were repeated using linear mixed-effects models with tissue class as a fixed effect and patient as a random intercept. The μ, SWS, μ-based dispersion, and joint metrics were log10-transformed for these models; omnibus effects were assessed by likelihood-ratio tests and pairwise contrasts by Bonferroni-corrected Wald tests. Patient-level cluster bootstrapping (2000 resamples) provided additional AUC and correlation confidence intervals. The nominal threshold was two-sided p < 0.05. Across 11 ttMRE tissue-class omnibus tests, Bonferroni-adjusted sensitivity analyses confirmed all 7 nominally significant findings (adjusted p = 0.002-0.027). Primary-tumor nodal-status and biomechanical-MRI association analyses were exploratory; the prespecified ttMRE μ-USE SWS correlation was assessed separately.

## 3. Results

### 3.1 Study cohort and sample inclusion

All 18 patients underwent in vivo USE before surgery. Ex vivo USE was available for 9 benign lymph nodes, 11 metastatic lymph nodes, and all 18 primary tumors (Fig. 1, Table 2). ttMRE was evaluable in 8 benign lymph nodes, 8 metastatic lymph nodes, and 13 primary tumors (7 node-negative, N-, and 6 node-positive, N+; Fig. 1, Table 1). ttMRE exclusions resulted from insufficient specimen dimensions (smaller than the glass tube diameter, 7 mm), inadequate wave-field quality, prioritization of tissue for diagnostic histopathology, or inability to complete the time-intensive measurement workflow. Of the 18 tumors initially diagnosed as HNSCC, one was reclassified as salivary duct carcinoma on final surgical histopathology. The salivary duct carcinoma showed biomechanical values within the range observed among the HNSCC tumors and was retained in the analysis; its exclusion did not alter the group-level conclusions.

**Table 2.** Ultrasound shear-wave elastography. SWS was measured before resection by in vivo USE in lymph nodes and after resection by ex vivo USE in nodes and primary tumors. Values are medians [IQR]; AUCs include 2000-resample bootstrap 95% confidence intervals. p values below 0.05 are bold. Abbreviations: LN, lymph node; N-, node-negative; N+, node-positive; SWS, shear-wave speed; AUC, area under the ROC curve; CI, confidence interval.

| Parameter | Benign LN | Metastatic LN | Primary tumor | Tumor N- | Tumor N+ | Group p | Benign vs meta | Benign vs tumor | Meta vs tumor | N- vs N+ p | AUC node detection [95% CI] | AUC nodal-status classification [95% CI] |
|---|---|---|---|---|---|---|---|---|---|---|---|---|
| SWS in vivo (m/s) | n = 9<br>1.92 [1.43-2.64] | n = 11<br>5.27 [4.29-5.69] | not imaged in vivo | - | - | <0.001 (Mann-Whitney) | <0.001 | - | - | - | 0.96 [0.86-1.00] | - |
| SWS ex vivo (m/s) | n = 9<br>3.32 [2.42-3.68] | n = 11<br>4.04 [3.63-4.39] | n = 18<br>4.17 [3.20-4.73] | n = 7<br>4.25 [4.17-4.68] | n = 11<br>3.24 [3.15-4.67] | 0.014 (Kruskal-Wallis) | 0.018 | 0.040 | 1.000 | 0.415 | 0.90 [0.75-1.00] | 0.62 [0.34-0.90] |

### 3.2 Cross-modal validation of tabletop MRE against ultrasound

ttMRE μ showed positive correlations with USE SWS (Fig. 2). Across 29 ex vivo specimens from 15 patients, ttMRE μ correlated with ex vivo USE SWS (Spearman ρ = 0.70, p < 0.001). For 16 lymph-node specimens from 14 patients that were also imaged before resection, ttMRE μ correlated with in vivo USE SWS (ρ = 0.69, p = 0.003); primary tumors were not examined by in vivo USE. These correlations support cross-modal rank-order agreement rather than equivalence of absolute values because USE reports m/s and ttMRE reports kPa under different loading and frequency conditions. Both associations were confirmed at the patient level (cluster-bootstrap 95% CI for ρ, 0.39-0.89 ex vivo and 0.30-0.86 in vivo; mixed-model p < 0.001 for each).

**Figure 2.**
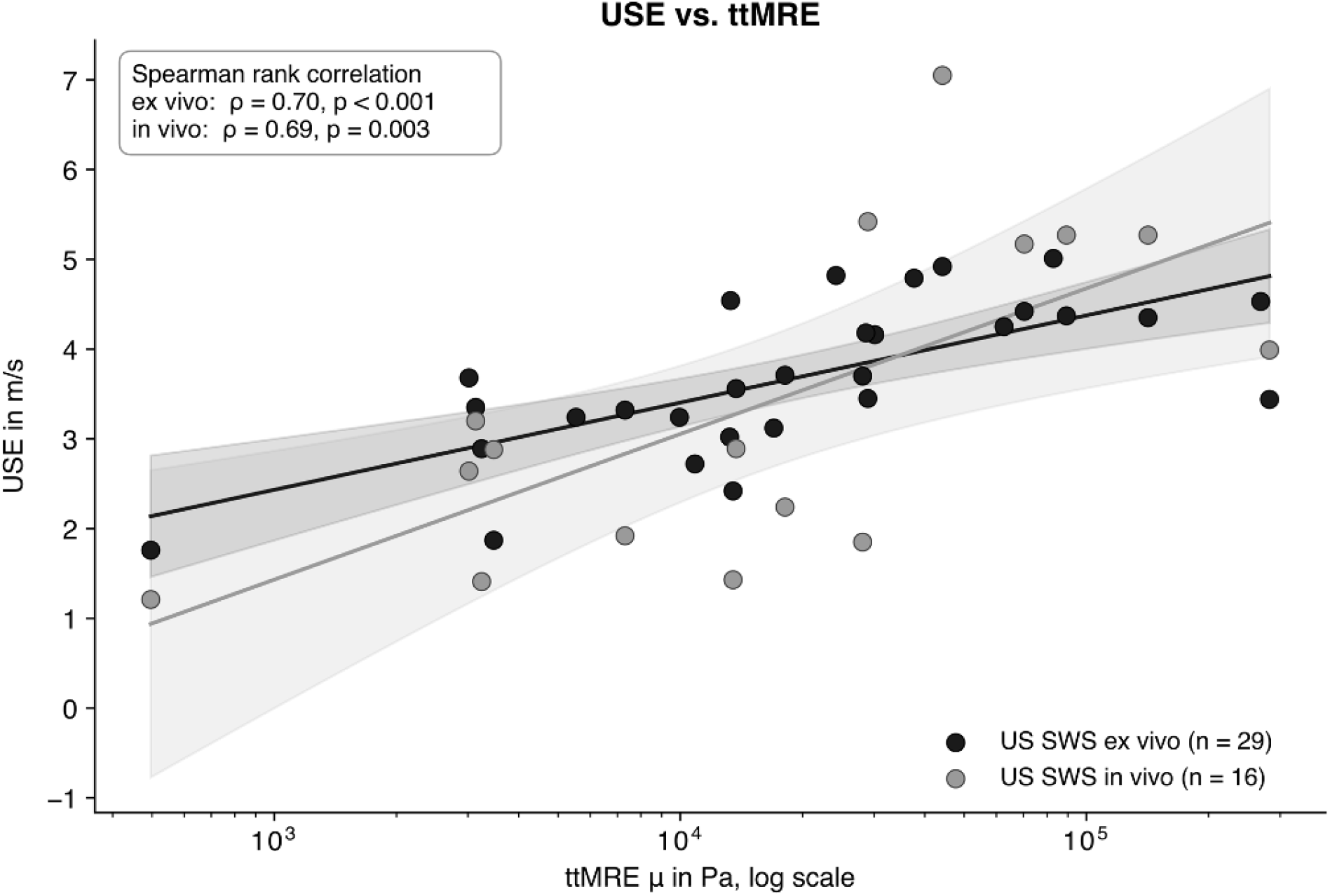
Agreement between ttMRE stiffness and USE shear-wave speed. USE shear-wave speed (SWS, m/s) is plotted against ttMRE μ on a logarithmic x axis. Black circles denote ex vivo USE (29 specimens from 15 patients; Spearman ρ = 0.70, p < 0.001), and gray circles denote in vivo USE of lymph nodes (16 specimens from 14 patients; ρ = 0.69, p = 0.003). Primary tumors were not examined in vivo. Two patients contributed both a benign and a metastatic node, so specimen counts exceed patient counts.

### 3.3 Stiffness discriminates benign from metastatic nodal tissue

Representative frequency-resolved wave fields and spring-pot fits (Fig. 3), and representative μ, α, fat-fraction, and ADC maps (Fig. 4) illustrate the analyzed data. ttMRE separated the tissue classes primarily through stiffness (Fig. 5, Table 1). Median μ was 3.4 kPa (IQR, 3.1-8.9) in benign lymph nodes, 57.3 kPa (26.2-102.4) in metastatic lymph nodes, and 24.2 kPa (13.2-37.6) in primary tumors (Kruskal-Wallis p = 0.001). Benign nodes differed from metastatic nodes (Dunn-Bonferroni p < 0.001) and primary tumors (p = 0.034), whereas metastatic nodes and primary tumors overlapped (p = 0.389). Ex vivo USE showed the same tissue-class pattern (Kruskal-Wallis p = 0.014): benign nodes had lower SWS than metastatic nodes (p = 0.018) and primary tumors (p = 0.040), while metastatic nodes and primary tumors did not differ (p = 1.000). In vivo USE also showed lower SWS in benign than metastatic nodes (Mann-Whitney p < 0.001). The overall α distributions did not differ significantly (Kruskal-Wallis p = 0.05). In the patient-level mixed-effects sensitivity analysis, the tissue-class differences were confirmed for ttMRE μ (p < 0.001; benign vs metastatic nodes p < 0.001, benign nodes vs primary tumors p = 0.001, metastatic nodes vs primary tumors p = 0.475) and ex vivo USE SWS (p < 0.001), and between-patient variance in μ was negligible. In this parametric model, α showed a nominal omnibus difference (p = 0.006) that was not present in any pairwise contrast (all p ≥ 0.37) and did not remain significant after correction across readouts (p = 0.068).

**Figure 3.**
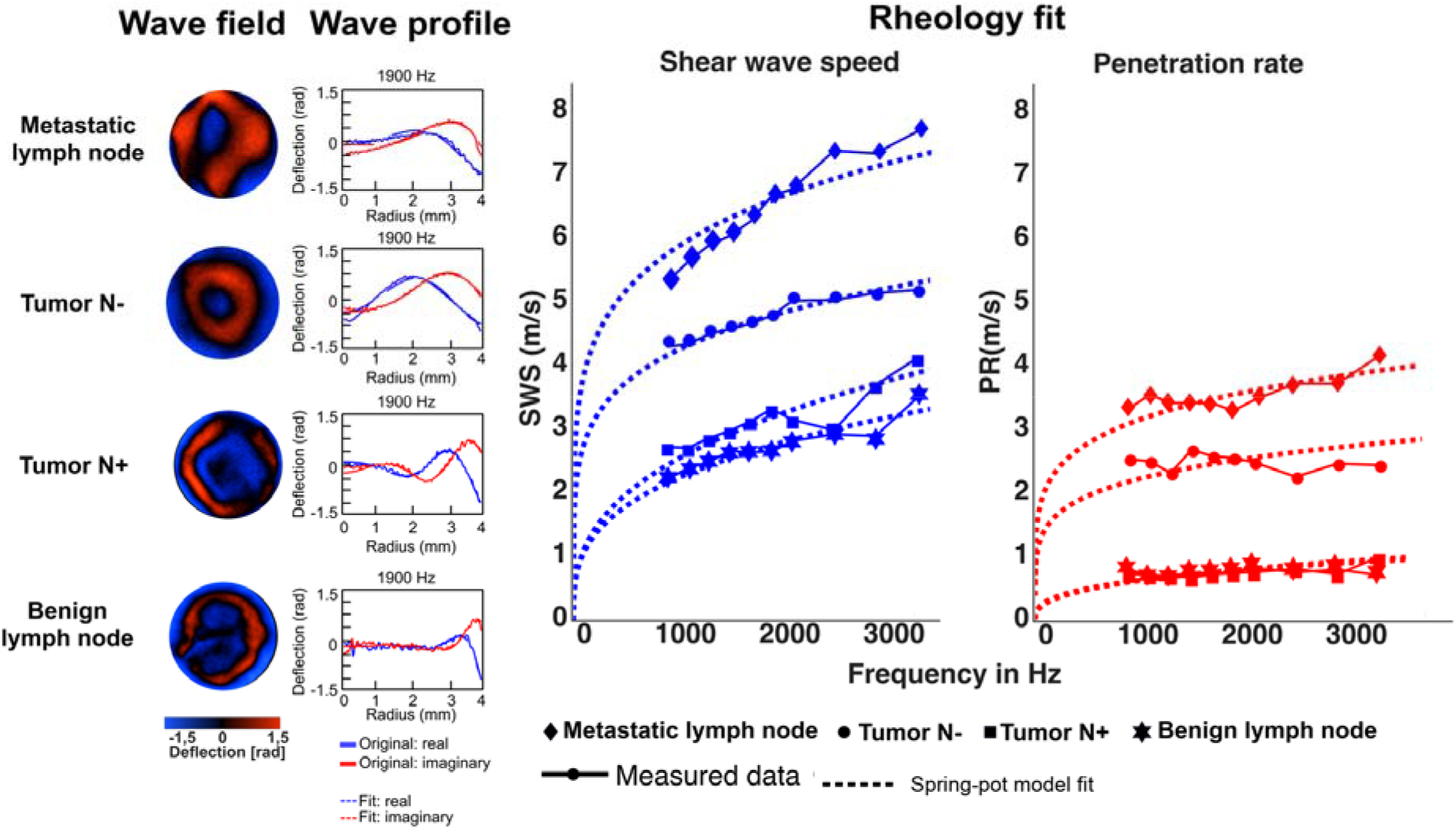
Complex wave profiles, wave fields, and spring-pot fits. Four specimens are shown: metastatic lymph node (diamond), node-negative primary tumor (circle), node-positive primary tumor (square), and benign lymph node (star). Left: wave-field images and complex radial wave profiles; blue and red denote the real and imaginary components, respectively. Axes show radius and complex wave-profile amplitude in arbitrary units. Right: frequency-resolved shear-wave speed (SWS, m/s) and penetration rate (PR, m/s) from 900 to 3300 Hz, with dotted spring-pot fits. Bessel fitting and spatial reconstruction are described in Section 2.4; group statistics appear in Table 1.

**Figure 4.**
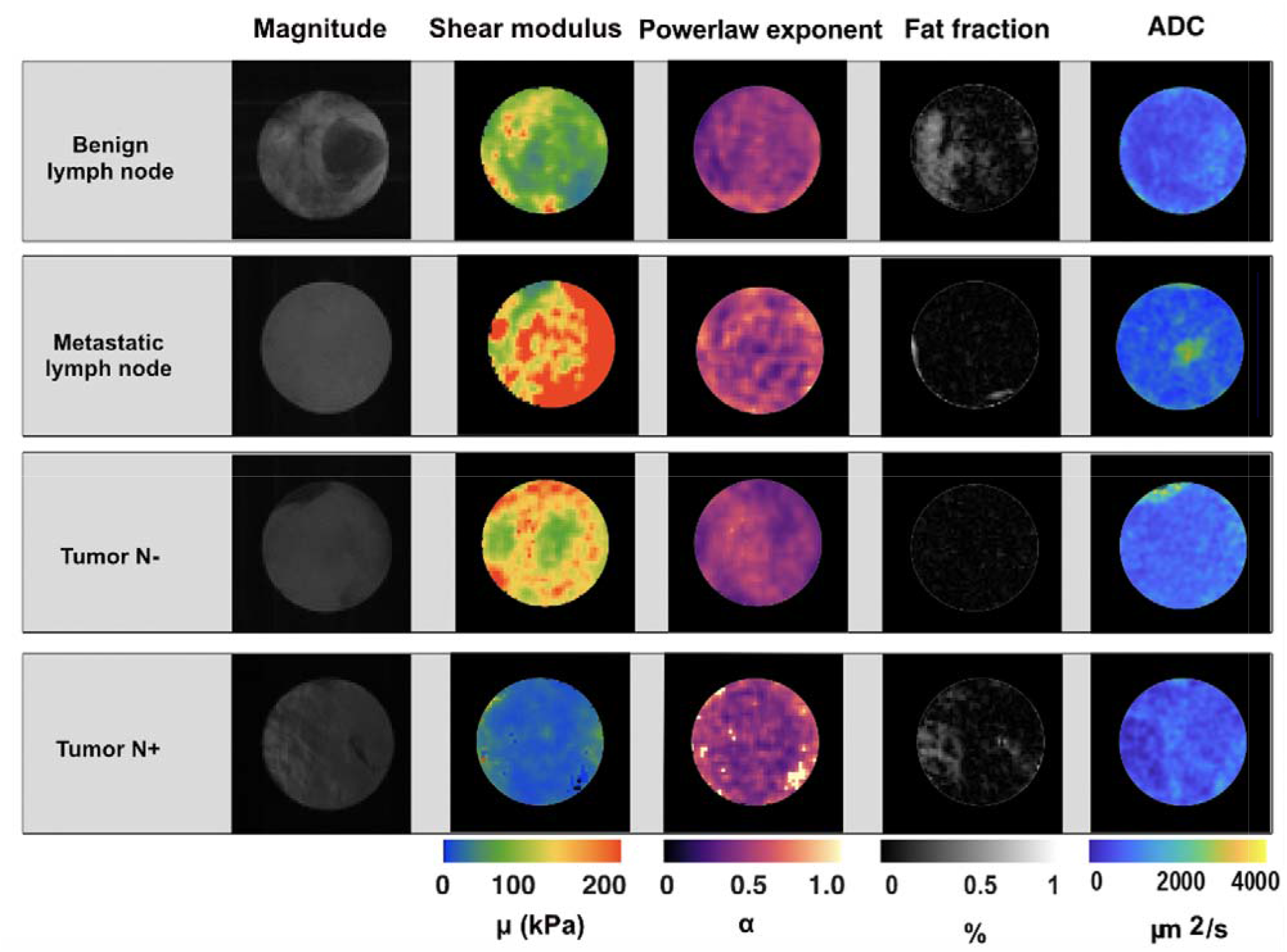
Representative biomechanical and multiparametric maps. Magnitude images and spatially resolved maps of shear modulus μ (kPa), dimensionless power-law exponent α, fat fraction (%), and apparent diffusion coefficient (ADC, µm²/s) are shown for a benign lymph node, metastatic lymph node, node-negative primary tumor, and node-positive primary tumor. α is defined on the interval 0-1. Illustrative specimens only; quantitative statistics are reported in Tables 1 and 3.

**Figure 5.**
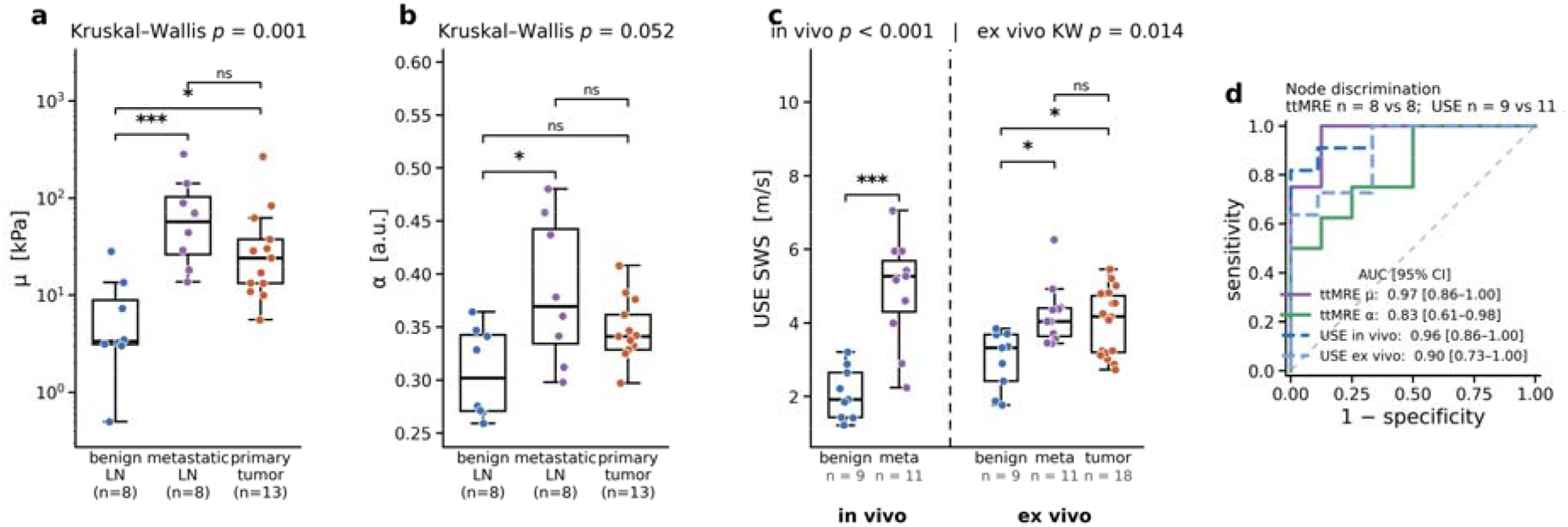
Discrimination of benign lymph nodes, metastatic lymph nodes, and primary tumors. (a, b) ttMRE μ and α acquired based on the rheological mapping in benign lymph nodes (n = 8), metastatic lymph nodes (n = 8), and primary tumors (n = 13). (c) In vivo USE shear-wave speed (SWS, m/s) in lymph nodes (benign n = 9; metastatic n = 11) and ex vivo USE SWS in benign lymph nodes (n = 9), metastatic lymph nodes (n = 11), and primary tumors (n = 18); the dashed vertical line separates the in vivo from the ex vivo cohort. In (a-c), brackets show Dunn-Bonferroni post-hoc tests for the three-group analyses and a Mann-Whitney U test for the in vivo USE comparison. (d) Receiver operating characteristic (ROC) analysis for discriminating metastatic from benign lymph nodes. Area under the curve (AUC) point estimates were 0.97 for ttMRE μ, 0.83 for ttMRE α, 0.96 for in vivo USE SWS, and 0.90 for ex vivo USE SWS; bootstrap confidence intervals are reported in Tables 1 and 2. μ is displayed on a logarithmic axis. *p < 0.05, **p < 0.01, ***p < 0.001; ns, not significant.

**Table 3.** Multiparametric MRI. T1, T2, fat fraction, and ADC were measured in sequence-specific ex vivo cohorts and compared among benign nodes, metastatic nodes, and primary tumors, with tumors stratified by nodal status. Values are medians [IQR]; AUCs include 2000-resample bootstrap 95% confidence intervals. p values below 0.05 are bold. Abbreviations: LN, lymph node; N-, node-negative; N+, node-positive; ADC, apparent diffusion coefficient; AUC, area under the ROC curve; CI, confidence interval.

| Parameter | Benign LN | Metastatic LN | Primary tumor | Tumor N- | Tumor N+ | KW<br>p | Benign vs meta | Benign vs tumor | Meta vs tumor | N- vs N+<br>p | AUC node detection<br>[95% CI] | AUC nodal-status classification<br>[95% CI] |
|---|---|---|---|---|---|---|---|---|---|---|---|---|
| T1 relaxation time (ms) | n = 8<br>562 [325-724] | n = 10<br>732 [679-838] | n = 15<br>858 [710-902] | n = 7<br>840 [747-881] | n = 8<br>860 [680-946] | 0.021 | 0.419 | 0.017 | 0.634 | 0.779 | 0.70 [0.45-0.93] | 0.55 [0.23-0.86] |
| T2 relaxation time (ms) | n = 8<br>125 [111-162] | n = 10<br>162 [103-166] | n = 15<br>131 [103-166] | n = 7<br>130 [103-144] | n = 8<br>142 [121-251] | 0.841 | 1.0 | 1.0 | 1.0 | 0.397 | 0.55 [0.25-0.85] | 0.64 [0.32-0.89] |
| Fat fraction (%) | n = 8<br>6.1 [2.2-53.9] | n = 10<br>2.0 [1.0-3.9] | n = 15<br>2.2 [0.6-4.0] | n = 7<br>0.8 [0.5-2.0] | n = 8<br>2.8 [1.9-5.6] | 0.09 | 0.227 | 0.109 | 1.0 | 0.152 | 0.75 [0.49-0.95] | 0.73 [0.45-0.96] |
| ADC (µm<sup>2</sup>/s) | n = 8<br>510 [450-650] | n = 10<br>480 [430-640] | n = 15<br>780 [730-860] | n = 7<br>780 [770-800] | n = 8<br>810 [680-880] | 0.005 | 1.000 | 0.062 | 0.010 | 0.955 | 0.52 [0.24-0.80] | 0.52 [0.21-0.82] |

### 3.4 Biomechanical heterogeneity of μ and α

Stiffness entropy H(μ) was lower in benign lymph nodes (median 1.24 nats, IQR 1.05-1.30) than in metastatic lymph nodes (1.88, 1.71-2.27) and primary tumors (1.70, 1.56-1.78; Kruskal-Wallis p = 0.002; Fig. 6, Table 1). SD(μ) and IQR(μ) also separated benign nodes from both malignant tissue classes (both omnibus p < 0.001), but these dispersion measures scale with μ and are not independent measures of heterogeneity. Joint IQR and joint SD perfectly separated metastatic from benign nodes in this dataset (AUC 1.00 for each), a sample-specific result that was numerically dominated by μ. Joint entropy H(μ,α) yielded an AUC of 0.92. None of the α-only heterogeneity measures showed a significant omnibus group difference (all p ≥ 0.090). These findings indicate that the apparent heterogeneity contrast was primarily stiffness related. The H(μ) difference was confirmed in the patient-level mixed model (p < 0.001).

**Figure 6.**
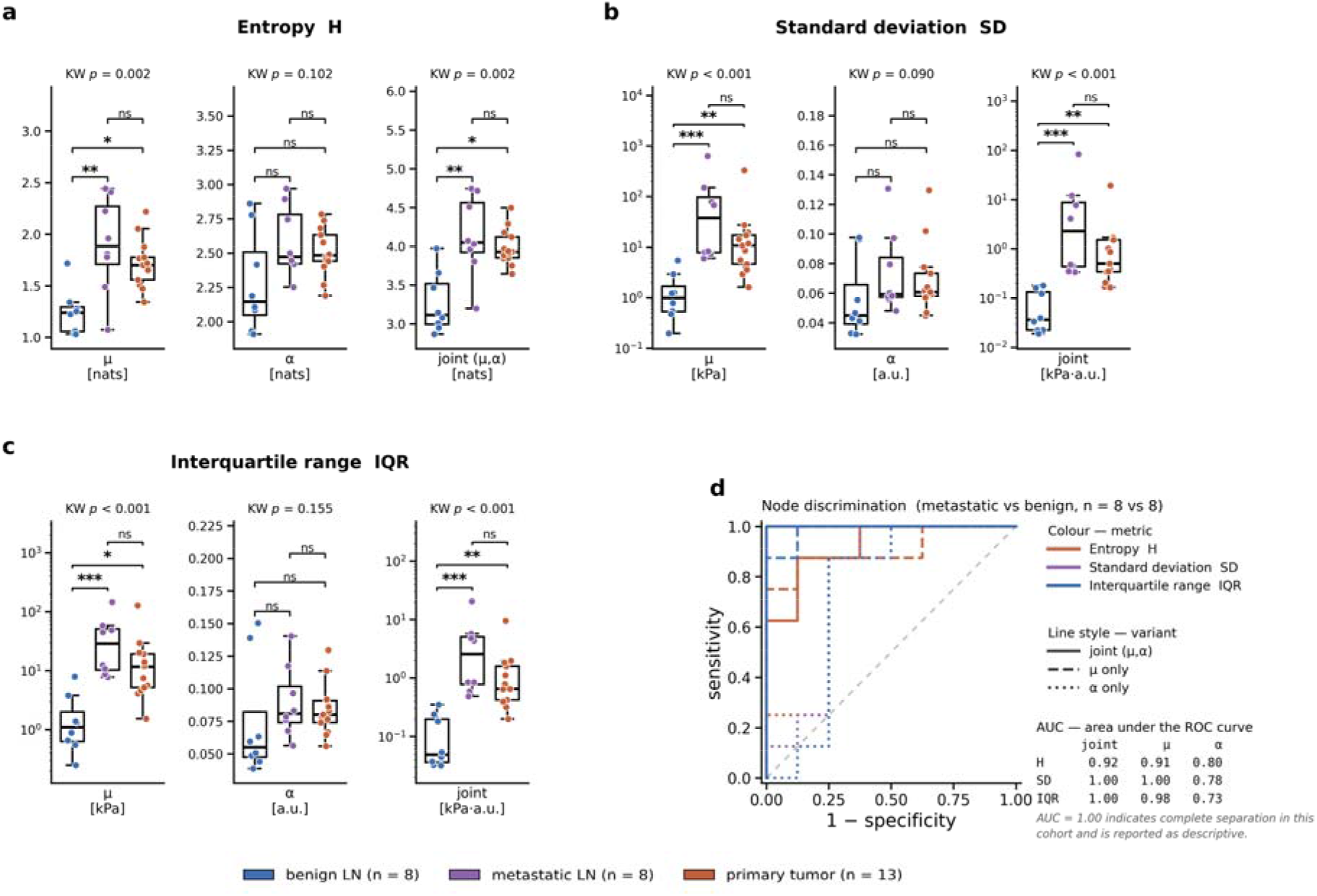
Biomechanical heterogeneity of μ and α. Panels show (a) Shannon entropy H, (b) within-specimen standard deviation SD, and (c) interquartile range IQR of μ and α in benign nodes (n = 8), metastatic nodes (n = 8), and primary tumors (n = 13). H(μ), H(α), and H(μ,α) indicate μ entropy, α entropy, and joint entropy (natural logarithm; nats). For each metric, μ-only, α-only, and joint μ-α values are plotted on separate y axes. μ-based and joint dispersion metrics use logarithmic axes in (b) and (c). Brackets show Dunn-Bonferroni contrasts. (d) ROC analysis for metastatic-versus-benign nodes (n = 8 each). Color denotes H, SD, or IQR; line style denotes joint (solid), μ-only (dashed), or α-only (dotted). AUCs (joint | μ | α) were 0.92 | 0.91 | 0.80 for H, 1.00 | 1.00 | 0.78 for SD, and 1.00 | 0.98 | 0.73 for IQR. Joint dispersion metrics were dominated by the μ term. Statistics are provided in Table 1. *p < 0.05, **p < 0.01, ***p < 0.001; ns, not significant.

### 3.5 Within-patient biomechanical differences

Matched comparisons clarified the relationship between the overlapping unpaired distributions of primary tumors and metastatic nodes (Fig. 7). In node-negative patients, primary tumors were stiffer than their paired benign nodes (median μ, 29.4 vs 3.4 kPa; Wilcoxon p = 0.031; n = 6 pairs); joint IQR and joint SD also increased (p = 0.031 for each), and ex vivo USE SWS showed the same direction (p = 0.047; n = 7 pairs). In node-positive patients, metastatic nodes were stiffer than their paired primary tumors (66.7 vs 12.1 kPa; p = 0.031; n = 6 pairs), whereas joint IQR, joint SD, and ex vivo USE SWS did not reach significance (all p ≥ 0.156). α and H(μ,α) were not significant at either step. At n = 6, p = 0.031 is the smallest attainable two-sided exact Wilcoxon p value and corresponds to concordant differences in all six pairs. These cross-sectional matched differences should not be interpreted as longitudinal progression.

**Figure 7.**
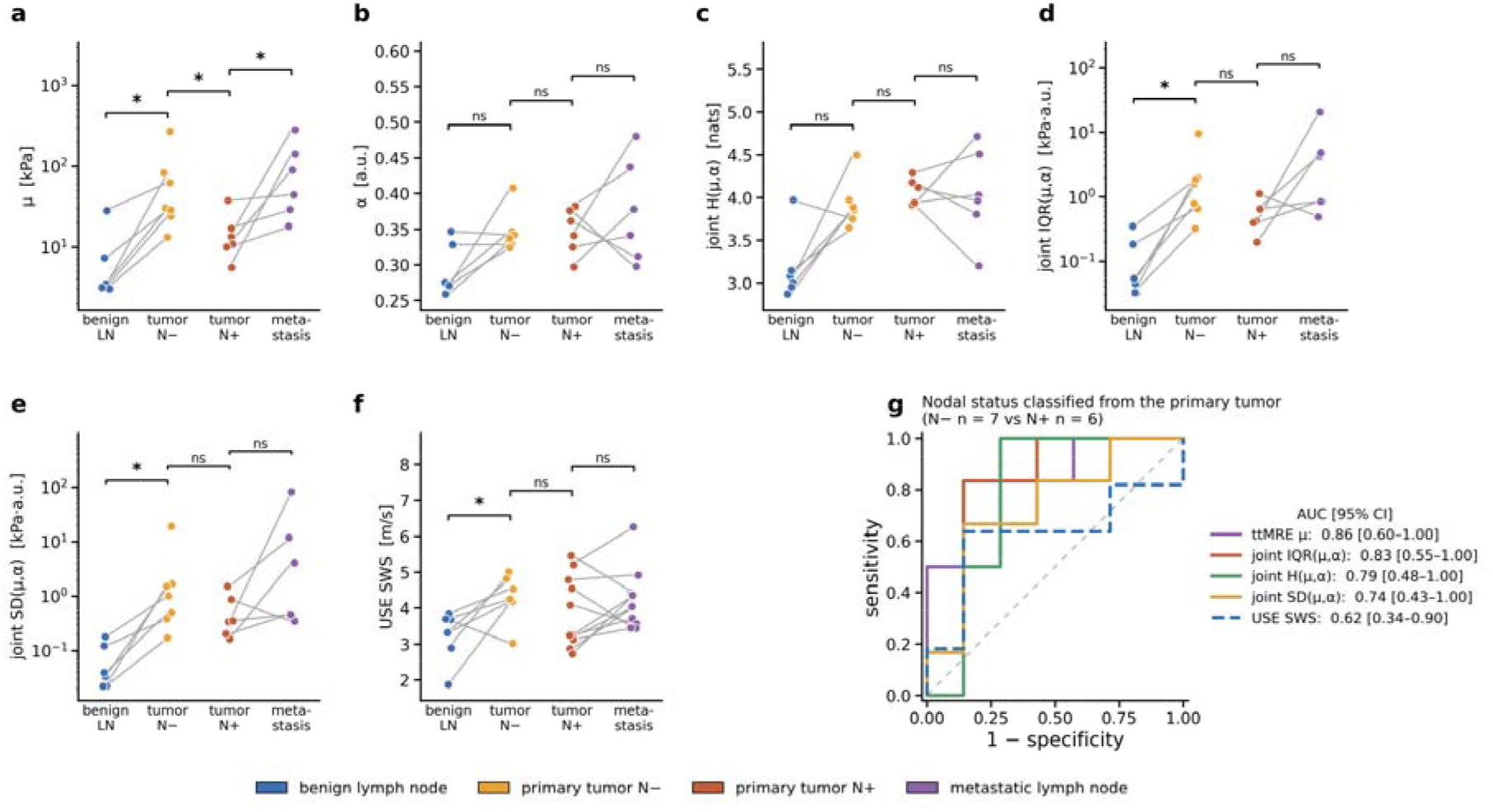
Within-patient biomechanical comparisons and exploratory classification of nodal status. (a-f) Gray lines link specimens from the same patient: benign node to node-negative tumor (ttMRE n = 6 pairs; ex vivo USE n = 7) and node-positive tumor to metastatic node (ttMRE n = 6 pairs; ex vivo USE n = 11). Panels show (a) μ, (b) α, (c) H(μ,α), (d) joint IQR, (e) joint SD, and (f) ex vivo USE SWS. Matched comparisons used Wilcoxon signed-rank tests; the unpaired node-negative-versus-node-positive comparison used Mann-Whitney U tests. The primary-tumor μ comparison yielded uncorrected p = 0.035. (g) ROC analysis of primary-tumor nodal status (ttMRE n = 7 node-negative and 6 node-positive; USE n = 7 and 11). AUCs were 0.86 for μ, 0.83 for joint IQR, 0.79 for H(μ,α), 0.74 for joint SD, and 0.62 for ex vivo USE SWS; confidence intervals are given in Tables 1 and 2. μ axes are logarithmic. *p < 0.05, **p < 0.01, ***p < 0.001; ns, not significant.

### 3.6 Primary-tumor stiffness by nodal status

In the secondary primary-tumor analysis, node-positive tumors had lower ttMRE μ than node-negative tumors (median 12.1 kPa [IQR, 10.2-16.1] vs 30.1 kPa [26.4-72.8]; two-sided Mann-Whitney uncorrected p = 0.035; Fig. 7, Table 1). The difference reached nominal significance but did not remain significant after correction across the 11 ttMRE readouts. Ex vivo USE SWS, available in all 18 tumors, did not differ by nodal status (p = 0.415; Table 2). α and H(μ,α) were also not significant (p = 0.836 and p = 0.101), while IQR(α) produced an isolated uncorrected p = 0.035. AUC point estimates for nodal status from the primary tumor were 0.86 for μ, 0.83 for joint IQR, 0.79 for H(μ,α), 0.74 for joint SD, and 0.62 for ex vivo USE SWS. The internally selected 17-kPa threshold for μ yielded 0.83 sensitivity and 0.86 specificity.

### 3.7 Node discrimination: tabletop MRE versus ultrasound

For discriminating metastatic from benign lymph nodes, AUC point estimates were 0.97 for ttMRE μ, 0.96 for in vivo USE SWS, 0.90 for ex vivo USE SWS, and 0.83 for ttMRE α (Fig. 5, Tables 1 and 2). Confidence intervals were wide and overlapping, and no formal comparisons among AUCs were performed. Internally selected Youden thresholds were 13.7 kPa for ttMRE μ (sensitivity 1.00, specificity 0.88), 3.99 m/s for in vivo USE (0.82, 1.00), and 3.44 m/s for ex vivo USE (1.00, 0.67). These operating points are descriptive and require external validation.

Patient-level cluster-bootstrap confidence intervals were similar to the specimen-level intervals (ttMRE μ, 0.85-1.00; ttMRE α, 0.60-1.00; in vivo USE, 0.84-1.00; ex vivo USE, 0.74-1.00).

### 3.8 Multiparametric MRI

Quantitative ex vivo MRI was available in sequence-specific cohorts (Table 3): T1, T2, and fat fraction in 8 benign lymph nodes, 10 metastatic nodes, and 15 primary tumors, and ADC in 8 benign lymph nodes, 10 metastatic nodes, and 15 primary tumors. T1 differed among tissue classes (Kruskal-Wallis p = 0.021): medians were 562 ms [IQR, 325-724] in benign nodes, 732 ms [679–838] in metastatic nodes, and 858 ms [710–902] in primary tumors. The benign-node-versus-primary-tumor comparison was significant (Dunn-Bonferroni p = 0.017). ADC also differed among tissue classes (Kruskal-Wallis p = 0.005), with lower values in metastatic nodes (480 [430–640] µm²/s) than in primary tumors (780 [730–860] µm²/s; p = 0.010); benign nodes were intermediate (510 [450–650] µm²/s) and did not differ from either group. T2 did not differ among groups. Fat fraction was higher and markedly more variable in benign nodes (6.1% [2.2-53.9]) than in metastatic nodes (2.0% [1.0-3.9]) and primary tumors (2.2% [0.6-4.0]), but the omnibus difference was not significant (p = 0.090). Across the 29 specimens with paired ttMRE and fat-fraction data, μ correlated inversely with fat fraction (Spearman ρ = -0.50, uncorrected p = 0.006). No quantitative MRI parameter differed by primary-tumor nodal status.

### 3.9 HPV/p16 status and matched tumor-node comparisons

Tumor μ, α, H(μ), and ex vivo USE SWS did not differ significantly by HPV/p16 status (all p > 0.4). Same-parameter correlations between a primary tumor and its matched node were also non-significant for μ, α, and H(μ) in both node-positive and node-negative subgroups (all p > 0.2; n = 6 per subgroup). These analyses were underpowered and are not interpreted as evidence of equivalence.

### 3.10 Qualitative histopathological context of the biomechanical phenotype

Representative histology provided qualitative context for the imaging findings (Fig. 8). Sections from a benign lymph node were compared with those from the three malignant tissue classes, namely a metastatic lymph node, a node-negative primary tumor, and a node-positive primary tumor. On H&E staining, all malignant specimens showed architectural disruption and an apparently denser and less uniformly distributed cellular infiltrate than benign nodal tissue. Sirius red staining showed more abundant collagen in the malignant specimens, with apparently thicker and more densely packed fiber bundles, and the OrientationJ fiber-orientation overlays indicated a visually more uniform fiber direction. Alcian blue staining revealed more extensive glycosaminoglycan-rich matrix, and MMP-9 immunoreactivity, used as a marker of extracellular-matrix degradation and remodeling, was more pronounced in the malignant specimens than in the benign lymph node. These observations are qualitative, are based on representative sections, and were not spatially registered to the ttMRE maps.

**Figure 8.**
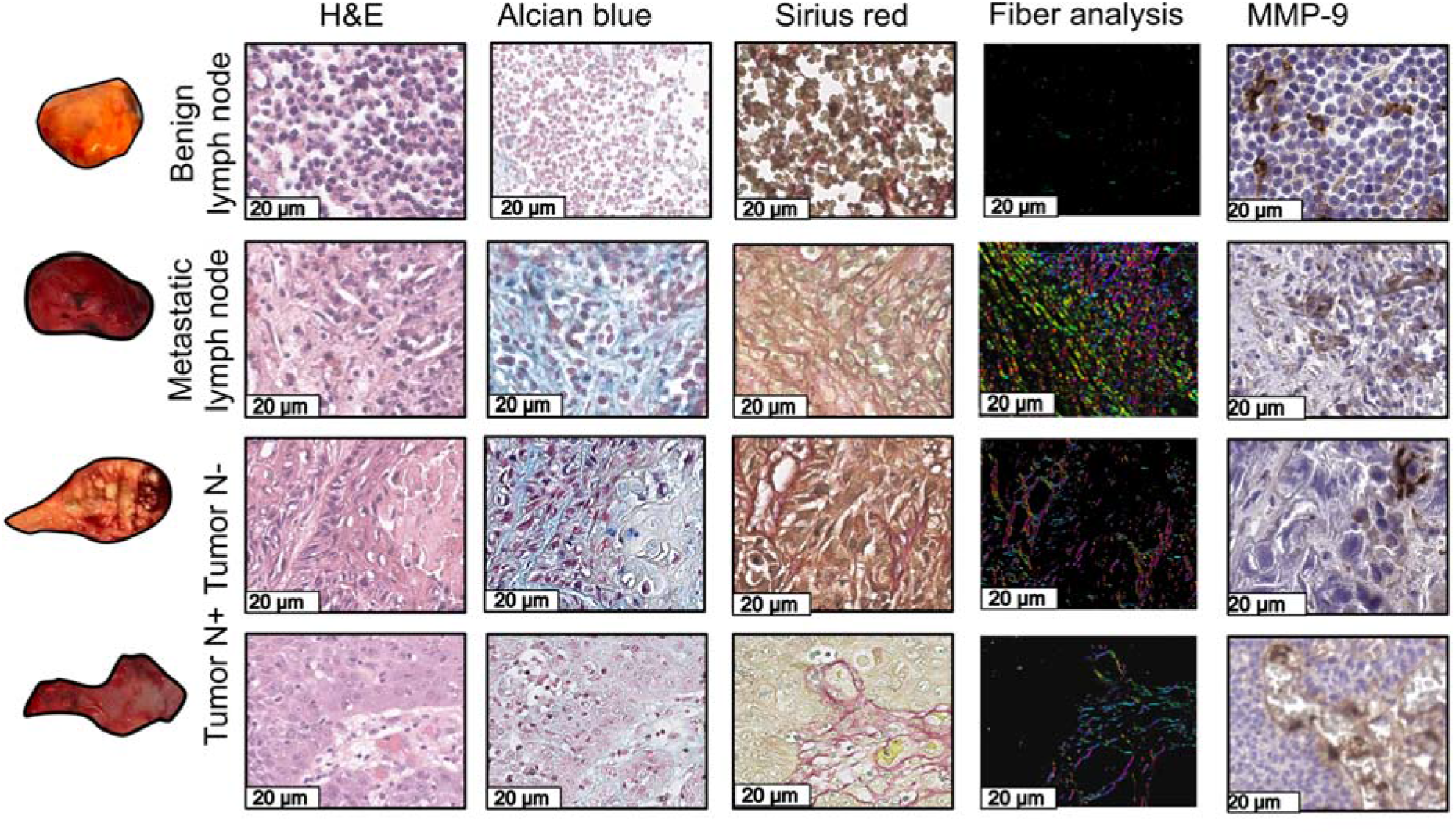
Representative histological sections providing qualitative context for the biomechanical findings. Macroscopic views and histological sections are shown for a benign lymph node, a metastatic lymph node, a node-negative primary tumor, and a node-positive primary tumor. Columns show hematoxylin and eosin (H&E) for tissue architecture and cellularity, Alcian blue for glycosaminoglycans (GAGs), Sirius red for collagen deposition and fiber thickness, a color-coded fiber-orientation overlay generated from the Sirius red sections with the OrientationJ plugin for ImageJ, and MMP-9 immunohistochemistry as a marker associated with extracellular-matrix degradation and remodeling. Qualitative findings are described in Section 3.10. Scale bars are 20 µm.

## 4. Discussion

Using in vivo and ex vivo USE together with spatially resolved multifrequency ttMRE, we characterized primary head and neck tumors and their corresponding cervical lymph nodes within the same patients. The principal finding was a consistent benign-versus-metastatic nodal stiffness contrast that was reproduced across all three acquisition settings and supported by positive cross-modal rank correlations. ttMRE additionally resolved stiffness-related spatial heterogeneity, although most heterogeneity measures remained strongly coupled to μ and therefore describe the same underlying contrast rather than an independent one. In a secondary analysis, primary tumors from node-positive patients had lower ttMRE μ than those from node-negative patients (uncorrected p = 0.035) - an exploratory association that was not reproduced by ex vivo USE and that points to a scale- and modality-dependent aspect of the tumor-node relationship.

### 4.1 Stiffness separates benign from malignant nodal tissue

Benign lymph nodes were consistently softer than metastatic nodes, in agreement with prior pathology-referenced cervical-node USE studies and with MRE-based nodal work [11, 12, 37, 38]. That this contrast persisted across in vivo USE, ex vivo USE, and ttMRE is notable, because these three settings differ substantially in excitation frequency, mechanical boundary conditions, and the physical quantity reported. A contrast that survives such differences is unlikely to be an artifact of any single acquisition geometry and more plausibly reflects a genuine difference in tissue composition. Among the heterogeneity metrics, H(μ) is less directly scale-dependent than SD(μ) and IQR(μ), since entropy responds to the shape of the parameter distribution rather than its absolute level. All three nevertheless remained closely related to μ and should be read as complementary descriptions of the same stiffness contrast rather than as independent confirmation of it [5].

### 4.2 Matched comparisons and patient-specific variation

The matched comparisons showed higher μ from benign node to primary tumor in node-negative patients, and from primary tumor to metastatic node in node-positive patients, with all six ttMRE pairs concordant at each step. Pairing removes between-patient baseline variation and can therefore expose consistent within-patient differences that remain hidden when unpaired group distributions overlap, as they did here. This interpretation has clear limits. The two paired comparisons involve different patient groups and cross-sectional specimens obtained at a single operation; they do not follow one lesion over time and consequently cannot demonstrate progressive mechanical amplification along a metastatic cascade. What they do support is a patient-specific mechanical context, in which the relevant quantity may be the stiffness difference between a patient’s own tissues rather than an absolute threshold applied across patients. Longitudinal designs and larger matched cohorts would be required to distinguish these possibilities.

### 4.3 Softer primary tumors are associated with nodal metastasis

Primary tumors from node-positive patients had lower stiffness by ttMRE than tumors from node-negative patients (uncorrected p = 0.035). Its interpretation depends critically on the scale of observation. Bulk tissue stiffness integrates extracellular matrix composition, cellularity, necrosis, fluid content, and measurement conditions, and is not equivalent to the stiffness of the individual cancer cells embedded within it. Mechanically soft or unjammed cancer-cell populations can coexist within an overall rigid tumor, and increased nuclear deformability has been shown to support invasion in HNSCC models [3, 52, 53]. A tumor that is softer at the tissue scale may therefore contain a larger fraction of deformable, migration-competent cells, or a matrix that has been partially degraded to permit their escape. These studies provide a plausible scale-dependent framework for the present association, but they do not establish its mechanism, which remains to be tested at the cellular level in this tumor entity.

### 4.4 Elastography discriminates metastatic nodes

The similar AUC estimates obtained with ttMRE μ and in vivo USE indicate that the nodal stiffness contrast is accessible with a clinically established modality, and not confined to a specialized ex vivo setup. Prior cervical-node studies have likewise reported higher USE stiffness in metastatic nodes [10–12], while MRE studies have described elevated image contrast in small abdominal nodal metastases and in cervical metastases assessed by DWI-based virtual elastography [37, 38]. The present design adds three elements to this literature: surgeon-guided retrieval of the specific node that was imaged, histopathological classification of that same node, and a direct in vivo-ex vivo comparison within one specimen. Other prospective in vivo cancer MRE studies have linked mechanical parameters to lymphovascular invasion, treatment response, and disease-free survival [27, 28]. Those results arise from different organs and different endpoints, however, and while they support the broader premise that tissue mechanics carry prognostic information, they do not validate the thresholds derived here.

### 4.5 Quantitative MRI findings

T1 was higher in primary tumors than in benign nodes, while ADC was lower in metastatic nodes than in primary tumors. Lower ADC in metastatic nodes is consistent with clinical diffusion-weighted imaging studies, in which metastatic cervical lymph nodes have consistently shown lower ADC than benign nodes [54, 55]; in the present ex vivo cohort, however, benign nodes were intermediate and did not differ from either group. These contrasts separate tissue classes other than the primary benign-versus-metastatic nodal comparison and did not classify the nodal status of primary tumors, indicating that the quantitative MRI parameters and the mechanical parameters respond to partly different tissue properties. The higher and more variable fat fraction observed in benign nodes plausibly reflects preserved fatty hila, a structure that is lost as a node becomes infiltrated; the same feature underlies the sonographic observation that a preserved echogenic hilum favors benignity, whereas loss of the hilum alone has limited specificity for malignancy [56]. Native T1 has been correlated with collagen area in an experimental hepatic tumor model [57], which offers a possible link between the T1 elevation seen here and matrix deposition, although no specimen-level correlation was performed in the present work.

Consistent with our observations, ex vivo parotid work reported higher T1 and lower fat fraction in pathological than in adjacent normal tissue, while mechanical parameters remained separate from an ADC-T1-T2 cluster [35], supporting the view that biomechanical and relaxometric contrasts provide partly complementary information.

### 4.6 Qualitative histopathological context for the biomechanical contrast

Representative histology showed several features with plausible but distinct biomechanical consequences (Fig. 8; schematically summarized in Fig. 9). First, architectural disruption together with a denser and less uniformly distributed cellularity could create local differences in cell packing and solid fraction, increasing the spatial variation of μ; the net effect on bulk stiffness is not uniquely predictable, however, because malignant cells themselves may be mechanically soft [3, 5]. Second, thicker and more densely packed collagen bundles would be expected to raise local matrix stiffness, while their more uniform orientation could produce anisotropic, direction-dependent shear responses; spatially uneven collagen deposition could therefore contribute to stiffness heterogeneity as much as to its absolute level [1, 2, 4]. Third, more extensive Alcian-blue-positive, glycosaminoglycan-rich matrix may alter water binding, swelling, permeability, and viscoelastic relaxation, and could thereby affect both μ and α [24, 26, 43]. Finally, stronger MMP-9 immunoreactivity is consistent with active extracellular-matrix turnover; the coexistence of collagen deposition and proteolytic degradation within the same specimen could generate adjacent stiffer and more fluid regions and so increase mechanical heterogeneity [26, 45]. These interpretations are offered as mechanistic hypotheses consistent with the observed contrasts, not as demonstrated relationships, since the histology was assessed qualitatively and was not spatially registered to the parameter maps.

**Figure 9.**
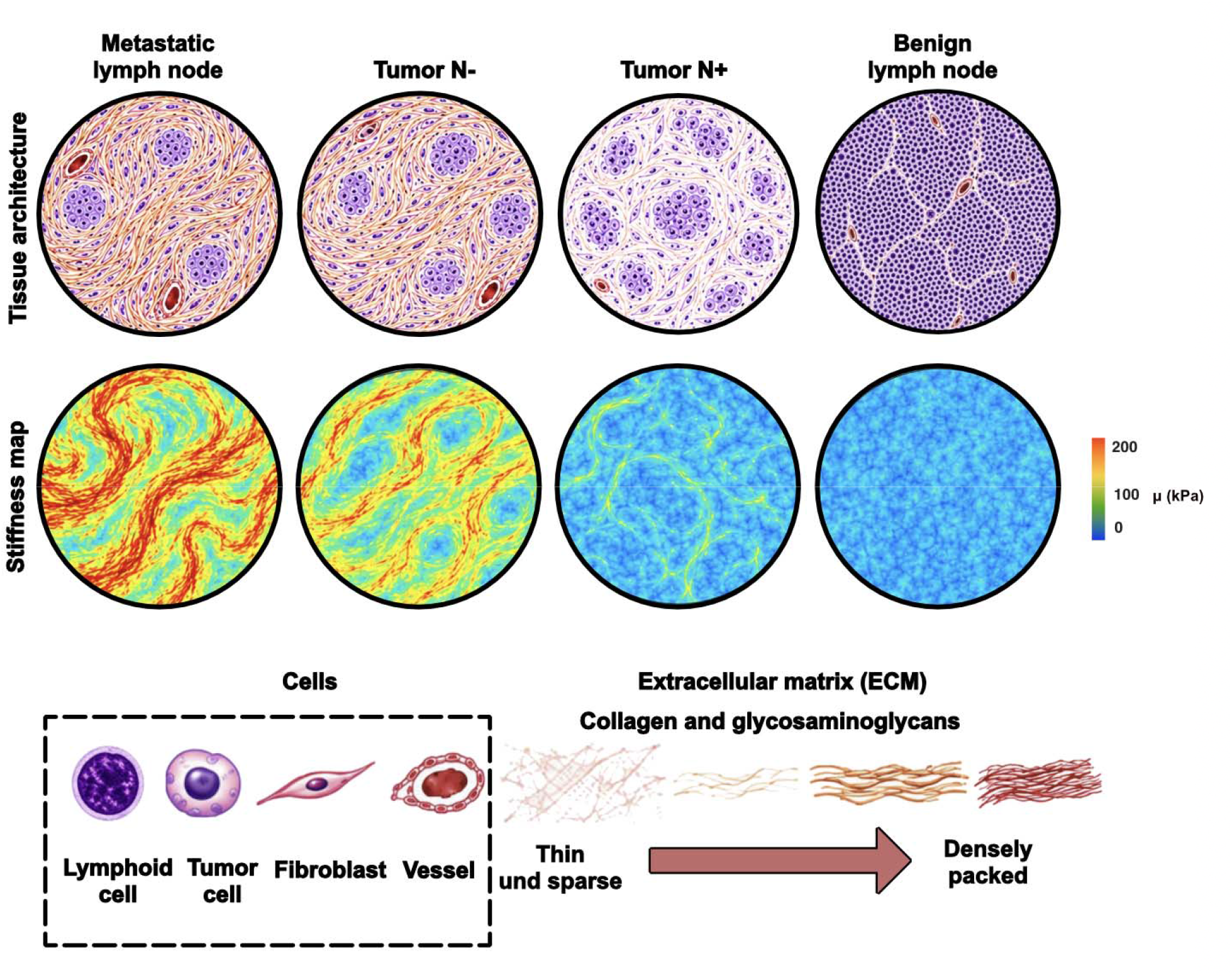
Schematic tissue architecture, relative stiffness, and extracellular-matrix organization. Conceptual illustration, not drawn to scale or derived from individual specimens, showing tissue architecture and relative stiffness for metastatic lymph nodes, node-negative primary tumors (Tumor N-), node-positive primary tumors (Tumor N+), and benign lymph nodes. Top: schematic cells, vessels, fibroblasts/cancer-associated fibroblasts (CAFs), and collagen organization. Bottom: schematic relative-stiffness maps. The ordering follows the cohort median ttMRE μ values: metastatic node 57.3 kPa, node-negative primary tumor 30.1 kPa, node-positive primary tumor 12.1 kPa, and benign node 3.4 kPa (Table 1). Colors represent relative, not absolute, stiffness.

### 4.7 Limitations

The cohort and matched subgroups were small, and one tumor was ultimately classified as salivary duct carcinoma rather than HNSCC. Not all scheduled specimens were evaluable because of specimen dimensions, wave quality or tissue allocation to diagnostic pathology. ttMRE and ex vivo USE differed from in vivo imaging in frequency and mechanical boundary conditions, so absolute values are not directly interchangeable. USE was performed by a single experienced examiner.

## 5. Conclusion

USE and ttMRE consistently distinguished benign from metastatic cervical nodes by stiffness, and ttMRE μ correlated with USE SWS. ttMRE also characterized stiffness-related spatial heterogeneity. Primary tumors from node-positive patients had lower ttMRE stiffness than those from node-negative patients, an exploratory association that was not seen with ex vivo USE. These findings provide a basis for further mechanical characterization of head and neck tumors and regional nodes.

## Data Availability

All data produced in the present study are available upon reasonable request to the authors

## Abbreviations

ADC: apparent diffusion coefficient
ARFI: acoustic radiation force impulse
AUC: area under the receiver-operating-characteristic curve
CI: confidence interval
G*: complex shear modulus
GAG: glycosaminoglycan
H: Shannon entropy (nats)
H&E: hematoxylin and eosin
H(μ), H(α), H(μ,α): Shannon entropy of μ, of α, and joint entropy of μ and α (nats)
HNSCC: head and neck squamous cell carcinoma
HPV: human papillomavirus
IQR: interquartile range
LN: lymph node
MMP-9: matrix metalloproteinase-9
MRE: magnetic resonance elastography
N- / N+: node-negative / node-positive
OrientationJ: ImageJ plugin used for color-coded collagen fiber-orientation overlays
PR: penetration rate
QuPath: whole-slide image analysis software
ROC: receiver operating characteristic
ROI: region of interest
SD: standard deviation
SP: spring-pot (fractional Scott-Blair) rheological model
SWS: shear-wave speed
ttMRE: tabletop magnetic resonance elastography
USE: ultrasound shear-wave elastography
G′: storage modulus (real part of the complex shear modulus)
G″: loss modulus (imaginary part of the complex shear modulus)
μ: shear modulus (stiffness) from the spring-pot model
α: power-law exponent from the spring-pot model (dimensionless)
φ: loss angle of the complex shear modulus
σ: residual of the global Bessel fit

## Author contributions

P. A. D. de Moraes: conceptualization, methodology, software, formal analysis, investigation, data curation, visualization, writing - original draft. M. Shahryari: methodology, writing - review and editing. S. Dommerich: investigation, resources, clinical data acquisition, writing - review and editing. H. Olze: resources, supervision, writing - review and editing. J. Pohl: investigation (histopathology), validation, writing - review and editing. I. Piwonski: investigation (histopathology), validation, writing - review and editing. T. Fischer: methodology (ultrasound elastography), validation, writing - review and editing. W. Zhao: methodology (tabletop MR elastography), software, writing - review and editing. K. Harb: investigation, data curation, writing - review and editing. J. Guo: methodology, software, formal analysis, writing - review and editing. I. Sack: conceptualization, methodology, resources, supervision, funding acquisition, writing - review and editing. K. M. Wakonig: conceptualization, investigation, project administration, supervision, writing - review and editing. All authors read and approved the final version of the manuscript.

## Acknowledgements

The authors thank the Institute of Pathology, Charité - Universitätsmedizin Berlin, for specimen processing and for the whole-slide digitalization of the histological sections. This work was supported by the German Research Foundation (Deutsche Forschungsgemeinschaft, DFG) through Collaborative Research Centre CRC 1340 “Matrix in Vision”, Research Unit FOR 5628 project 513752256, and the Research Training Group BIOQIC (GRK 2260).

## Data availability

Specimen-level frequency-resolved parameters are provided in Supplementary Table S3, with retained frequencies and exclusion reasons in Supplementary Table S1 and group summaries in Supplementary Table S2, and in the accompanying data workbook. Further data are available from the corresponding author on reasonable request.

