## Supplementary material for "Biomechanical characterization of head and neck tumors and corresponding lymph nodes for nodal assessment by ultrasound shear wave elastography and MR elastography"

**Contents**

- Supplementary Methods S1 — Frequency-resolved reconstruction and derived quantities
- Supplementary Methods S2 — Frequency retention and quality criteria
- Supplementary Methods S3 — Spring-pot model and its relation to the reported quantities
- Supplementary Methods S4 — Heterogeneity metrics: definitions and scale dependence
- Supplementary Methods S5 — Qualitative histological assessment
- Supplementary Table S1 — Retained frequencies and exclusion reasons per specimen
- Supplementary Table S2 — Frequency-resolved group summary
- Supplementary Table S3 — Specimen-level frequency-resolved parameters (complete listing)
- Supplementary Table S4 — Nomenclature and symbol conventions

**Supplementary Methods S1 — Frequency-resolved reconstruction and derived quantities**

At each drive frequency f, the complex wave field was obtained after temporal Fourier transformation of the four phase offsets and spatial phase unwrapping. A global Bessel-function fit to the complex radial wave profile provided a specimen-level complex wavenumber k* = k′ + ik″ and served as a prior for directional bandpass filtering. Two real-valued, frequency-resolved quantities were reconstructed from the spatial phase and amplitude gradients of the filtered fields [1–4].

Shear-wave speed was calculated as

$SWS(f) = 2\pi f / k'(f)$	(S1)

SWS is reported in m/s and increases with tissue stiffness and, in dispersive media, with frequency.

Penetration rate was calculated as

$PR(f) = 2\pi f / k''(f)$	(S2)

PR is reported in m/s, expresses reciprocal spatial attenuation as a velocity, and decreases as viscous damping increases.

The symbols SWS and PR are used consistently throughout the manuscript, all figures and tables, the data workbook and this Supplement.

From the same complex wavenumber, the specimen-level complex shear modulus was obtained as

$G^{*}(f) = \rho[2\pi f / k^{*}(f)]^{2}$	(S3)

where tissue density was assumed to be ρ = 1000 kg/m³. The storage modulus G′, loss modulus G″, and loss angle φ were defined as

$G' = |G^{*}| cos \varphi; G'' = |G^{*}| sin \varphi; \varphi= arctan(G''/G')$	(S4)

Here G′ and G″ are the real and imaginary parts of G*, and φ = 0 represents a purely elastic material and φ = π/2 a purely viscous material. G′ and G″ are reported in kPa and φ in radians. The Bessel-fit residual σ is a dimensionless goodness-of-fit measure of the global complex-wave-profile fit; lower values indicate closer agreement between the measured radial profile and the Bessel model.

**Supplementary Methods S2 — Frequency retention and quality criteria**

The actuator was driven across a nominal range of 800 to 5000 Hz (four steps); 800 Hz did not yield evaluable wave fields in any specimen, so retained frequencies span 900–5000 Hz. A frequency was retained only when the shear-wave amplitude and phase signal-to-noise ratio remained above the noise floor across the central analysis region of the 7-mm tube, and when the wavelength was sufficiently sampled by the 0.15-mm pixel grid to permit reliable estimation of the spatial phase gradient.

Three exclusion categories were recorded. Low-frequency exclusion denotes drive frequencies at which no propagating shear wave could be established in the specimen. Long-wavelength exclusion denotes frequencies at which the wavelength exceeded the specimen dimensions, so that less than approximately one wavelength was contained in the field of view. Specimens smaller than the 7-mm tube diameter, or for which no tissue remained after diagnostic histopathology, were excluded before measurement.

Across the 29 specimens of the final analysis cohort, 269 specimen-frequency combinations were retained. Retained frequencies spanned 900–5000 Hz; in most specimens the usable range was 900–3300 Hz, with a median of 10 retained frequencies per specimen (range 5–10 across the 28 specimens with an evaluable specimen-level fit; see below for the one specimen without). The nominal extremes of the drive range were rarely usable: 800 Hz was retained in no specimen, and 4700–5000 Hz only in the stiffest metastatic lymph node. Supplementary Table S1 lists the retained frequencies and the exclusion reason for every specimen.

**Supplementary Methods S3 — Spring-pot model and its relation to the reported quantities**

The frequency-resolved SWS and PR values at each pixel were fitted directly with a two-parameter fractional spring-pot model [1–5], defined as

$G^{*}(\omega) = \mu^{1-\alpha}\left( \frac{i\omega}{\omega_{0}} \right)^{\alpha},$	(S5)

where G* is the complex shear modulus, ω is angular frequency, and ω₀ = 2πf₀ is the reference angular frequency with f₀ = 1000 Hz. The parameter μ is the shear modulus at the reference frequency (reported in kPa), and the dimensionless power-law exponent 0 ≤ α ≤ 1 describes the transition from elastic-like (α → 0) to viscous-like (α → 1) behavior. Because one pair (μ, α) is fitted to all retained frequencies at each pixel, μ and α are frequency-independent summary parameters, whereas SWS, PR, G′, G″, and φ are frequency-resolved.

Two levels of analysis are therefore reported. The pixel-wise spring-pot maps yield μ and α and all heterogeneity metrics, and form the basis of every group comparison in the main manuscript. The specimen-level values tabulated in Supplementary Tables S2 and S3 derive from the global Bessel fit and characterize the specimen as a whole. The two levels are complementary and are not numerically identical; specimen-level values are provided for transparency and for comparison with other tabletop MRE implementations, and were not used for the statistics reported in the manuscript. One specimen, the benign lymph node of patient 18, contributed no evaluable frequency at the specimen level while retaining a valid pixel-wise spring-pot fit; it is therefore listed in Supplementary Table S1 without frequency-resolved values but was included in all group statistics.

**Supplementary Methods S4 — Heterogeneity metrics: definitions and scale dependence**

| **Metric** | **Definition** | **Scale behavior** |
| --- | --- | --- |
| SD | within-specimen standard deviation of the pixel values | scales with the mean; increases with the μ level |
| IQR | difference between the 75th and 25th percentiles of the pixel values | scales with the mean; robust to outliers |
| H (Shannon entropy) | Equation (S6), evaluated over a 30-bin histogram; H is reported in nats | scale-free; depends on distribution shape rather than level |
| H(μ,α) | joint entropy from the 30 × 30 two-dimensional histogram of paired μ–α pixels, in nats | scale-free; weights both axes independently |
| joint SD, joint IQR | products SD(μ) × SD(α) and IQR(μ) × IQR(α) | numerically dominated by the μ term |

For normalized histogram probabilities pᵢ, Shannon entropy was calculated as described in the main manuscript [6]:

$H = -\Sigmaᵢ pᵢ ln(pᵢ)$	(S6)

Because μ spanned several orders of magnitude, its histogram bin edges were defined in log₁₀ space from the pooled 0.5th–99.5th percentiles, whereas α was binned linearly. The use of log₁₀ for bin placement does not change the natural-logarithm base in Equation (S6). Univariate normalized entropy was calculated as H/ln(30).

Quantitative consequence. Separating the joint products into their components shows that the discriminative information resides almost entirely in μ. For metastatic-versus-benign node discrimination (n = 8 vs 8), the AUCs of the μ-only and joint variants were nearly identical (entropy, 0.91 vs 0.92; SD, 1.00 vs 1.00; IQR, 0.98 vs 1.00), whereas the α-only variants performed substantially worse (0.80, 0.78 and 0.73) and reached significance in no Kruskal–Wallis test (p = 0.102, 0.090 and 0.155). Joint metrics should therefore be interpreted as μ-weighted descriptors rather than as independent measures of μ–α coupling.

**Supplementary Methods S5 — Qualitative histological assessment**

Whole-slide images were digitized at the Institute of Pathology, Charité – Universitätsmedizin Berlin, and reviewed in QuPath v0.4.3 [7]. Assessment was qualitative throughout: sections were evaluated by visual inspection, and no image-based measurements, area fractions, densities, or optical-density values were extracted. The features listed below were therefore recorded as descriptive impressions rather than as numerical variables, and none of them entered any statistical analysis.

| **Stain / marker** | **Feature assessed visually** | **Biomechanical interpretation** |
| --- | --- | --- |
| H&E | tissue architecture; apparent density and spatial uniformity of the cellular infiltrate | cellular packing; qualitative counterpart of the imaging-derived heterogeneity |
| Alcian blue | apparent extent of GAG-rich matrix | water-binding, poroelastic matrix component; contributes to fluidity and to the spatial variance of μ |
| Sirius red | apparent abundance of collagen and thickness of fiber bundles | principal load-bearing constituent; primary determinant of μ |
| Sirius red + OrientationJ overlay (ImageJ) | predominant fiber direction and visual uniformity of fiber alignment (illustrative overlay only) | fiber alignment and structural order of the matrix |
| MMP-9 immunohistochemistry | apparent staining intensity and extent | extracellular-matrix degradation and remodeling |

For illustration, color-coded fiber-orientation overlays of the Sirius red sections were generated with the OrientationJ plugin for ImageJ [8]. These overlays display the local predominant fiber direction as a colour map and were used only to make differences in the visual uniformity of fiber direction apparent; no orientation statistics, coherency values, or angular dispersion measures were computed from them. Histological evaluation was not registered pixel-by-pixel to the ttMRE maps, and no specimen-level imaging–histology correlation was performed.

**Supplementary Table S1 — Retained frequencies and exclusion reasons per specimen**

*Final analysis cohort of 29 specimens. Frequencies are drive frequencies of the tabletop MRE actuator.*

| **Specimen** | **Tissue** | **n** | **Range [Hz]** | **Retained frequencies [Hz]** | **Exclusion reason** |
| --- | --- | --- | --- | --- | --- |
| mpHNSCC-003 benign LN | Benign LN | 10 | 900–3300 | 900, 1100, 1300, 1500, 1700, 1900, 2100, 2500, 2900, 3300 | — |
| mpHNSCC-004 benign LN | Benign LN | 10 | 900–3300 | 900, 1100, 1300, 1500, 1700, 1900, 2100, 2500, 2900, 3300 | — |
| mpHNSCC-005 benign LN | Benign LN | 10 | 900–3300 | 900, 1100, 1300, 1500, 1700, 1900, 2100, 2500, 2900, 3300 | — |
| mpHNSCC-006 benign LN | Benign LN | 8 | 900–2500 | 900, 1100, 1300, 1500, 1700, 1900, 2100, 2500 | — |
| mpHNSCC-011 benign LN | Benign LN | 10 | 900–3300 | 900, 1100, 1300, 1500, 1700, 1900, 2100, 2500, 2900, 3300 | — |
| mpHNSCC-015 benign LN | Benign LN | 10 | 900–3300 | 900, 1100, 1300, 1500, 1700, 1900, 2100, 2500, 2900, 3300 | — |
| mpHNSCC-017 benign LN | Benign LN | 10 | 900–3300 | 900, 1100, 1300, 1500, 1700, 1900, 2100, 2500, 2900, 3300 | — |
| mpHNSCC-018 benign LN | Benign LN | 0 | — | — | low-frequency exclusion (no propagating shear wave) |
| mpHNSCC-001 metastatic LN | Metastatic LN | 10 | 900–3300 | 900, 1100, 1300, 1500, 1700, 1900, 2100, 2500, 2900, 3300 | — |
| mpHNSCC-002 metastatic LN | Metastatic LN | 10 | 900–3300 | 900, 1100, 1300, 1500, 1700, 1900, 2100, 2500, 2900, 3300 | — |
| mpHNSCC-009 metastatic LN | Metastatic LN | 10 | 900–3300 | 900, 1100, 1300, 1500, 1700, 1900, 2100, 2500, 2900, 3300 | — |
| mpHNSCC-012 metastatic LN | Metastatic LN | 10 | 900–3300 | 900, 1100, 1300, 1500, 1700, 1900, 2100, 2500, 2900, 3300 | — |
| mpHNSCC-014 metastatic LN | Metastatic LN | 8 | 1300–3300 | 1300, 1500, 1700, 1900, 2100, 2500, 2900, 3300 | long-wavelength exclusion (wavelength exceeds specimen) |
| mpHNSCC-016 metastatic LN | Metastatic LN | 10 | 900–3300 | 900, 1100, 1300, 1500, 1700, 1900, 2100, 2500, 2900, 3300 | — |
| mpHNSCC-017 metastatic LN | Metastatic LN | 10 | 900–3300 | 900, 1100, 1300, 1500, 1700, 1900, 2100, 2500, 2900, 3300 | — |
| mpHNSCC-018 metastatic LN | Metastatic LN | 9 | 1500–5000 | 1500, 1700, 1900, 2100, 2500, 2900, 3300, 4700, 5000 | long-wavelength exclusion (wavelength exceeds specimen) |
| mpHNSCC-001 tumor | Primary tumor | 10 | 900–3300 | 900, 1100, 1300, 1500, 1700, 1900, 2100, 2500, 2900, 3300 | — |
| mpHNSCC-002 tumor | Primary tumor | 10 | 900–3300 | 900, 1100, 1300, 1500, 1700, 1900, 2100, 2500, 2900, 3300 | — |
| mpHNSCC-003 tumor | Primary tumor | 10 | 900–3300 | 900, 1100, 1300, 1500, 1700, 1900, 2100, 2500, 2900, 3300 | — |
| mpHNSCC-004 tumor | Primary tumor | 10 | 900–3300 | 900, 1100, 1300, 1500, 1700, 1900, 2100, 2500, 2900, 3300 | — |
| mpHNSCC-005 tumor | Primary tumor | 10 | 900–3300 | 900, 1100, 1300, 1500, 1700, 1900, 2100, 2500, 2900, 3300 | — |
| mpHNSCC-006 tumor | Primary tumor | 10 | 900–3300 | 900, 1100, 1300, 1500, 1700, 1900, 2100, 2500, 2900, 3300 | — |
| mpHNSCC-011 tumor | Primary tumor | 10 | 900–3300 | 900, 1100, 1300, 1500, 1700, 1900, 2100, 2500, 2900, 3300 | — |
| mpHNSCC-013 tumor | Primary tumor | 5 | 1900–3300 | 1900, 2100, 2500, 2900, 3300 | long-wavelength exclusion (wavelength exceeds specimen) |
| mpHNSCC-014 tumor | Primary tumor | 10 | 900–3300 | 900, 1100, 1300, 1500, 1700, 1900, 2100, 2500, 2900, 3300 | — |
| mpHNSCC-015 tumor | Primary tumor | 10 | 900–3300 | 900, 1100, 1300, 1500, 1700, 1900, 2100, 2500, 2900, 3300 | — |
| mpHNSCC-016 tumor | Primary tumor | 9 | 900–2900 | 900, 1100, 1300, 1500, 1700, 1900, 2100, 2500, 2900 | long-wavelength exclusion (wavelength exceeds specimen) |
| mpHNSCC-017 tumor | Primary tumor | 10 | 900–3300 | 900, 1100, 1300, 1500, 1700, 1900, 2100, 2500, 2900, 3300 | — |
| mpHNSCC-018 tumor | Primary tumor | 10 | 900–3300 | 900, 1100, 1300, 1500, 1700, 1900, 2100, 2500, 2900, 3300 | — |

**Supplementary Table S2 — Frequency-resolved group summary**

*Median [interquartile range] per tissue class and drive frequency; n is the number of specimens contributing a value at that frequency. G′ and G″ in kPa, φ in radians, σ dimensionless.*

| **Tissue** | **f [Hz]** | **n** | **SWS [m/s]** | **PR [m/s]** | **G′ [kPa]** | **G″ [kPa]** | **φ [rad]** | **σ** |
| --- | --- | --- | --- | --- | --- | --- | --- | --- |
| Benign LN | 900 | 7 | 2.73 [2.18–3.08] | 1.00 [0.80–1.26] | 3.29 [2.99–6.09] | 4.80 [2.88–5.60] | 0.77 [0.69–0.89] | 0.54 [0.43–0.62] |
| Benign LN | 1100 | 7 | 2.71 [2.22–3.15] | 0.94 [0.74–1.29] | 3.18 [2.54–6.40] | 4.77 [2.97–5.87] | 0.76 [0.72–0.90] | 0.42 [0.34–0.48] |
| Benign LN | 1300 | 7 | 2.87 [2.45–3.24] | 0.95 [0.74–1.33] | 3.41 [2.65–6.77] | 5.12 [3.71–6.29] | 0.75 [0.73–1.02] | 0.29 [0.24–0.36] |
| Benign LN | 1500 | 7 | 2.98 [2.59–3.29] | 0.86 [0.80–1.31] | 3.35 [3.03–6.84] | 5.35 [4.26–6.75] | 0.81 [0.76–1.01] | 0.26 [0.22–0.32] |
| Benign LN | 1700 | 7 | 3.13 [2.68–3.44] | 0.80 [0.73–1.39] | 3.11 [2.53–7.46] | 5.89 [4.58–7.40] | 0.96 [0.75–1.08] | 0.24 [0.21–0.30] |
| Benign LN | 1900 | 7 | 3.32 [2.75–3.57] | 0.77 [0.76–1.26] | 2.76 [2.60–6.82] | 7.01 [4.78–7.96] | 0.95 [0.82–1.19] | 0.22 [0.21–0.30] |
| Benign LN | 2100 | 7 | 3.16 [2.87–3.67] | 0.90 [0.86–1.26] | 3.99 [3.51–6.99] | 6.47 [5.18–8.53] | 0.86 [0.84–1.09] | 0.15 [0.14–0.22] |
| Benign LN | 2500 | 7 | 2.99 [2.67–3.70] | 1.09 [0.72–1.30] | 4.71 [2.52–7.61] | 5.79 [4.40–8.67] | 0.86 [0.84–1.11] | 0.09 [0.09–0.13] |
| Benign LN | 2900 | 6 | 3.58 [3.31–4.05] | 0.94 [0.76–1.32] | 4.02 [2.67–8.11] | 7.67 [6.15–10.34] | 0.91 [0.90–1.17] | 0.11 [0.09–0.12] |
| Benign LN | 3300 | 6 | 4.18 [3.33–4.47] | 0.93 [0.77–1.21] | 3.84 [2.39–7.31] | 7.67 [5.46–12.40] | 0.98 [0.92–1.15] | 0.13 [0.12–0.16] |
| Metastatic LN | 900 | 6 | 4.65 [4.13–5.02] | 1.90 [1.46–2.48] | 12.45 [8.53–18.94] | 10.39 [9.00–14.91] | 0.76 [0.58–0.79] | 0.52 [0.46–0.60] |
| Metastatic LN | 1100 | 6 | 4.98 [4.30–5.83] | 1.94 [1.63–2.30] | 15.00 [11.72–19.20] | 14.72 [9.56–21.47] | 0.81 [0.75–0.86] | 0.42 [0.36–0.56] |
| Metastatic LN | 1300 | 7 | 5.76 [4.46–6.69] | 1.79 [1.44–2.20] | 15.34 [8.52–18.38] | 18.40 [11.31–29.07] | 0.89 [0.76–1.01] | 0.34 [0.26–0.39] |
| Metastatic LN | 1500 | 8 | 6.03 [4.58–7.18] | 1.82 [1.51–2.55] | 13.43 [8.35–18.79] | 21.53 [12.27–31.06] | 0.87 [0.77–1.11] | 0.31 [0.28–0.38] |
| Metastatic LN | 1700 | 8 | 5.88 [4.64–6.98] | 2.08 [1.75–2.38] | 16.96 [14.69–22.71] | 19.87 [12.39–31.02] | 0.88 [0.78–0.97] | 0.29 [0.25–0.33] |
| Metastatic LN | 1900 | 8 | 6.28 [4.78–7.01] | 1.91 [1.76–2.35] | 17.44 [14.40–20.01] | 24.09 [13.76–31.88] | 0.97 [0.80–1.05] | 0.28 [0.26–0.34] |
| Metastatic LN | 2100 | 8 | 6.12 [4.82–6.37] | 2.05 [1.55–2.47] | 15.81 [11.34–20.35] | 22.81 [12.32–25.99] | 0.89 [0.71–1.06] | 0.23 [0.20–0.26] |
| Metastatic LN | 2500 | 8 | 5.74 [5.20–6.19] | 2.12 [1.51–2.59] | 18.06 [11.20–26.21] | 17.10 [13.47–24.81] | 0.95 [0.66–1.07] | 0.16 [0.10–0.18] |
| Metastatic LN | 2900 | 8 | 5.59 [5.12–6.58] | 2.10 [1.60–2.49] | 18.35 [11.82–23.40] | 18.74 [15.52–27.18] | 0.89 [0.79–1.02] | 0.12 [0.10–0.20] |
| Metastatic LN | 3300 | 8 | 5.69 [5.06–6.84] | 2.04 [1.47–2.35] | 17.10 [10.33–23.17] | 20.12 [14.95–29.65] | 0.93 [0.82–1.03] | 0.15 [0.12–0.22] |
| Metastatic LN | 4700 | 1 | 7.91 [7.91–7.91] | 2.47 [2.47–2.47] | 29.20 [29.20–29.20] | 40.20 [40.20–40.20] | 0.94 [0.94–0.94] | 0.09 [0.09–0.09] |
| Metastatic LN | 5000 | 1 | 7.86 [7.86–7.86] | 2.45 [2.45–2.45] | 28.76 [28.76–28.76] | 39.66 [39.66–39.66] | 0.94 [0.94–0.94] | 0.08 [0.08–0.08] |
| Primary tumor | 900 | 12 | 3.97 [3.28–5.61] | 1.63 [1.27–1.96] | 8.28 [7.29–16.26] | 9.96 [5.95–20.03] | 0.83 [0.66–0.92] | 0.44 [0.37–0.47] |
| Primary tumor | 1100 | 12 | 4.08 [3.39–5.33] | 1.64 [1.28–2.09] | 8.92 [7.54–16.14] | 10.32 [6.23–17.36] | 0.80 [0.68–0.94] | 0.34 [0.30–0.44] |
| Primary tumor | 1300 | 12 | 4.22 [3.43–5.29] | 1.66 [1.23–1.87] | 9.31 [7.53–14.09] | 11.04 [6.84–16.69] | 0.87 [0.68–1.00] | 0.26 [0.25–0.37] |
| Primary tumor | 1500 | 12 | 4.41 [3.46–5.21] | 1.64 [1.36–1.86] | 10.42 [8.13–13.47] | 12.06 [7.21–17.34] | 0.88 [0.67–0.96] | 0.26 [0.24–0.35] |
| Primary tumor | 1700 | 12 | 4.47 [3.52–5.34] | 1.65 [1.24–1.97] | 10.54 [7.57–13.97] | 12.30 [7.25–17.86] | 0.80 [0.71–0.97] | 0.24 [0.22–0.30] |
| Primary tumor | 1900 | 13 | 4.63 [3.75–5.47] | 1.62 [1.45–2.18] | 10.85 [9.31–15.47] | 13.60 [9.14–19.27] | 0.88 [0.69–1.02] | 0.26 [0.25–0.32] |
| Primary tumor | 2100 | 13 | 4.63 [4.00–5.58] | 1.69 [1.40–2.22] | 11.59 [9.54–14.89] | 13.21 [9.71–19.90] | 0.83 [0.78–1.05] | 0.18 [0.14–0.20] |
| Primary tumor | 2500 | 13 | 5.03 [3.80–5.98] | 1.59 [1.44–2.31] | 11.11 [10.23–12.03] | 16.19 [7.94–23.04] | 0.88 [0.79–1.03] | 0.11 [0.09–0.14] |
| Primary tumor | 2900 | 13 | 5.79 [4.19–7.52] | 1.64 [1.47–3.16] | 12.98 [9.59–37.08] | 20.38 [10.95–32.77] | 0.88 [0.72–1.00] | 0.11 [0.10–0.15] |
| Primary tumor | 3300 | 12 | 5.34 [4.41–6.71] | 1.68 [1.44–2.51] | 12.93 [9.34–21.73] | 18.18 [11.90–26.97] | 0.89 [0.80–1.02] | 0.12 [0.11–0.19] |

**Supplementary Table S3 — Specimen-level frequency-resolved parameters**

*Complete listing of every evaluable specimen-frequency combination in the final analysis cohort (269 rows). SWS, shear-wave speed; PR, penetration rate; G′, storage modulus; G″, loss modulus; φ, loss angle; σ, residual of the global Bessel fit. Values derive from the global Bessel fit and are complementary to the pixel-wise spring-pot parameters reported in the manuscript.*

| **Specimen** | **N status** | **f [Hz]** | **SWS [m/s]** | **PR [m/s]** | **G′ [kPa]** | **G″ [kPa]** | **φ [rad]** | **σ** |
| --- | --- | --- | --- | --- | --- | --- | --- | --- |
| mpHNSCC-003 benign LN | — | 900 | 2.139 | 1.005 | 3.258 | 2.494 | 0.653 | 0.542 |
| mpHNSCC-003 benign LN | — | 1100 | 2.188 | 0.936 | 3.183 | 2.748 | 0.712 | 0.417 |
| mpHNSCC-003 benign LN | — | 1300 | 2.295 | 0.950 | 3.408 | 3.075 | 0.734 | 0.268 |
| mpHNSCC-003 benign LN | — | 1500 | 2.309 | 0.862 | 3.123 | 3.255 | 0.806 | 0.245 |
| mpHNSCC-003 benign LN | — | 1700 | 2.356 | 0.718 | 2.494 | 3.577 | 0.962 | 0.193 |
| mpHNSCC-003 benign LN | — | 1900 | 2.446 | 0.758 | 2.757 | 3.849 | 0.949 | 0.211 |
| mpHNSCC-003 benign LN | — | 2100 | 2.583 | 0.891 | 3.572 | 4.186 | 0.864 | 0.151 |
| mpHNSCC-003 benign LN | — | 2500 | 2.745 | 1.094 | 4.712 | 4.474 | 0.760 | 0.093 |
| mpHNSCC-003 benign LN | — | 2900 | 3.460 | 0.547 | 0.042 | 5.944 | 1.564 | 0.122 |
| mpHNSCC-003 benign LN | — | 3300 | 4.222 | 0.458 | 2.066 | 5.266 | 1.197 | 0.131 |
| mpHNSCC-004 benign LN | — | 900 | 2.727 | 0.823 | 3.286 | 4.801 | 0.971 | 0.617 |
| mpHNSCC-004 benign LN | — | 1100 | 2.714 | 0.695 | 2.356 | 4.766 | 1.112 | 0.403 |
| mpHNSCC-004 benign LN | — | 1300 | 2.867 | 0.684 | 2.184 | 5.252 | 1.177 | 0.294 |
| mpHNSCC-004 benign LN | — | 1500 | 2.985 | 0.774 | 2.931 | 5.770 | 1.101 | 0.259 |
| mpHNSCC-004 benign LN | — | 1700 | 3.133 | 0.743 | 2.563 | 6.261 | 1.182 | 0.237 |
| mpHNSCC-004 benign LN | — | 1900 | 3.324 | 0.772 | 2.716 | 7.013 | 1.201 | 0.213 |
| mpHNSCC-004 benign LN | — | 2100 | 3.157 | 0.837 | 3.449 | 6.466 | 1.081 | 0.183 |
| mpHNSCC-004 benign LN | — | 2500 | 2.992 | 0.771 | 2.902 | 5.793 | 1.106 | 0.132 |
| mpHNSCC-004 benign LN | — | 2900 | 3.704 | 0.812 | 2.784 | 8.544 | 1.256 | 0.148 |
| mpHNSCC-004 benign LN | — | 3300 | 4.130 | 0.724 | 0.898 | 9.303 | 1.475 | 0.173 |
| mpHNSCC-005 benign LN | — | 900 | 2.039 | 0.651 | 2.005 | 2.660 | 0.925 | 0.246 |
| mpHNSCC-005 benign LN | — | 1100 | 2.077 | 0.648 | 2.010 | 2.770 | 0.943 | 0.228 |
| mpHNSCC-005 benign LN | — | 1300 | 2.107 | 0.553 | 1.501 | 2.879 | 1.090 | 0.199 |
| mpHNSCC-005 benign LN | — | 1500 | 2.125 | 0.607 | 1.815 | 2.929 | 1.016 | 0.179 |
| mpHNSCC-005 benign LN | — | 1700 | 2.240 | 0.603 | 1.790 | 3.259 | 1.068 | 0.144 |
| mpHNSCC-005 benign LN | — | 1900 | 2.372 | 0.559 | 1.444 | 3.585 | 1.188 | 0.172 |
| mpHNSCC-005 benign LN | — | 2100 | 2.466 | 0.643 | 2.023 | 3.942 | 1.097 | 0.133 |
| mpHNSCC-005 benign LN | — | 2500 | 2.588 | 0.664 | 2.145 | 4.334 | 1.111 | 0.094 |
| mpHNSCC-005 benign LN | — | 2900 | 2.304 | 0.749 | 2.625 | 3.383 | 0.911 | 0.102 |
| mpHNSCC-005 benign LN | — | 3300 | 2.339 | 0.914 | 3.359 | 3.278 | 0.773 | 0.133 |
| mpHNSCC-006 benign LN | — | 900 | 2.227 | 0.785 | 2.723 | 3.090 | 0.848 | 0.529 |
| mpHNSCC-006 benign LN | — | 1100 | 2.249 | 0.778 | 2.716 | 3.169 | 0.862 | 0.495 |
| mpHNSCC-006 benign LN | — | 1300 | 2.597 | 0.805 | 3.108 | 4.337 | 0.949 | 0.423 |
| mpHNSCC-006 benign LN | — | 1500 | 2.873 | 0.825 | 3.350 | 5.354 | 1.012 | 0.365 |
| mpHNSCC-006 benign LN | — | 1700 | 3.014 | 0.796 | 3.114 | 5.893 | 1.085 | 0.343 |
| mpHNSCC-006 benign LN | — | 1900 | 3.350 | 0.753 | 2.486 | 7.051 | 1.232 | 0.322 |
| mpHNSCC-006 benign LN | — | 2100 | 3.457 | 0.903 | 3.993 | 7.746 | 1.095 | 0.285 |
| mpHNSCC-006 benign LN | — | 2500 | 0.615 | 0.131 | 0.070 | 0.233 | 1.280 | 0.265 |
| mpHNSCC-011 benign LN | — | 900 | 2.872 | 1.122 | 5.060 | 4.943 | 0.774 | 0.334 |
| mpHNSCC-011 benign LN | — | 1100 | 2.904 | 1.224 | 5.537 | 4.878 | 0.722 | 0.269 |
| mpHNSCC-011 benign LN | — | 1300 | 2.966 | 1.236 | 5.722 | 5.116 | 0.730 | 0.217 |
| mpHNSCC-011 benign LN | — | 1500 | 2.992 | 1.214 | 5.690 | 5.274 | 0.747 | 0.204 |
| mpHNSCC-011 benign LN | — | 1700 | 3.129 | 1.359 | 6.590 | 5.576 | 0.702 | 0.236 |
| mpHNSCC-011 benign LN | — | 1900 | 3.063 | 1.149 | 5.525 | 5.717 | 0.803 | 0.215 |
| mpHNSCC-011 benign LN | — | 2100 | 3.147 | 1.110 | 5.442 | 6.171 | 0.848 | 0.132 |
| mpHNSCC-011 benign LN | — | 2500 | 3.225 | 1.117 | 5.595 | 6.516 | 0.861 | 0.085 |
| mpHNSCC-011 benign LN | — | 2900 | 3.264 | 1.060 | 5.263 | 6.787 | 0.911 | 0.084 |
| mpHNSCC-011 benign LN | — | 3300 | 3.064 | 0.948 | 4.320 | 6.039 | 0.950 | 0.082 |
| mpHNSCC-015 benign LN | — | 900 | 4.930 | 2.362 | 17.538 | 13.101 | 0.642 | 0.926 |
| mpHNSCC-015 benign LN | — | 1100 | 4.915 | 2.218 | 16.730 | 13.477 | 0.678 | 0.792 |
| mpHNSCC-015 benign LN | — | 1300 | 5.073 | 2.379 | 18.309 | 14.047 | 0.654 | 0.407 |
| mpHNSCC-015 benign LN | — | 1500 | 4.982 | 2.565 | 18.706 | 12.787 | 0.600 | 0.468 |
| mpHNSCC-015 benign LN | — | 1700 | 5.112 | 2.683 | 19.900 | 13.295 | 0.589 | 0.399 |
| mpHNSCC-015 benign LN | — | 1900 | 4.811 | 2.443 | 17.305 | 12.026 | 0.607 | 0.422 |
| mpHNSCC-015 benign LN | — | 2100 | 4.807 | 1.944 | 14.642 | 13.638 | 0.750 | 0.250 |
| mpHNSCC-015 benign LN | — | 2500 | 5.245 | 1.858 | 15.197 | 17.115 | 0.845 | 0.125 |
| mpHNSCC-015 benign LN | — | 2900 | 5.564 | 1.832 | 15.588 | 19.664 | 0.901 | 0.120 |
| mpHNSCC-015 benign LN | — | 3300 | 5.689 | 1.862 | 16.167 | 20.591 | 0.905 | 0.209 |
| mpHNSCC-017 benign LN | — | 900 | 3.290 | 1.390 | 7.122 | 6.252 | 0.720 | 0.620 |
| mpHNSCC-017 benign LN | — | 1100 | 3.403 | 1.361 | 7.265 | 6.871 | 0.758 | 0.458 |
| mpHNSCC-017 benign LN | — | 1300 | 3.519 | 1.418 | 7.821 | 7.319 | 0.752 | 0.305 |
| mpHNSCC-017 benign LN | — | 1500 | 3.597 | 1.414 | 7.988 | 7.733 | 0.769 | 0.279 |
| mpHNSCC-017 benign LN | — | 1700 | 3.746 | 1.415 | 8.322 | 8.531 | 0.798 | 0.248 |
| mpHNSCC-017 benign LN | — | 1900 | 3.787 | 1.369 | 8.115 | 8.860 | 0.829 | 0.286 |
| mpHNSCC-017 benign LN | — | 2100 | 3.883 | 1.404 | 8.535 | 9.316 | 0.829 | 0.138 |
| mpHNSCC-017 benign LN | — | 2500 | 4.172 | 1.480 | 9.631 | 10.824 | 0.844 | 0.083 |
| mpHNSCC-017 benign LN | — | 2900 | 4.165 | 1.410 | 9.065 | 10.942 | 0.879 | 0.065 |
| mpHNSCC-017 benign LN | — | 3300 | 4.550 | 1.300 | 8.311 | 13.431 | 1.017 | 0.115 |
| mpHNSCC-001 metastatic LN | — | 900 | 5.742 | 2.248 | 20.270 | 19.747 | 0.772 | 0.377 |
| mpHNSCC-001 metastatic LN | — | 1100 | 5.606 | 2.352 | 20.564 | 18.223 | 0.725 | 0.350 |
| mpHNSCC-001 metastatic LN | — | 1300 | 5.762 | 2.631 | 23.193 | 18.404 | 0.671 | 0.288 |
| mpHNSCC-001 metastatic LN | — | 1500 | 5.721 | 2.894 | 24.416 | 17.054 | 0.610 | 0.272 |
| mpHNSCC-001 metastatic LN | — | 1700 | 5.859 | 3.096 | 26.238 | 17.385 | 0.585 | 0.211 |
| mpHNSCC-001 metastatic LN | — | 1900 | 5.975 | 2.673 | 24.566 | 20.014 | 0.684 | 0.206 |
| mpHNSCC-001 metastatic LN | — | 2100 | 6.031 | 2.478 | 23.375 | 21.301 | 0.739 | 0.151 |
| mpHNSCC-001 metastatic LN | — | 2500 | 5.739 | 3.023 | 25.133 | 16.711 | 0.587 | 0.084 |
| mpHNSCC-001 metastatic LN | — | 2900 | 6.019 | 2.695 | 24.949 | 20.303 | 0.683 | 0.092 |
| mpHNSCC-001 metastatic LN | — | 3300 | 6.180 | 2.336 | 22.664 | 23.203 | 0.797 | 0.096 |
| mpHNSCC-002 metastatic LN | — | 900 | 4.076 | 1.555 | 9.953 | 10.059 | 0.791 | 0.580 |
| mpHNSCC-002 metastatic LN | — | 1100 | 4.282 | 1.588 | 10.668 | 11.220 | 0.811 | 0.470 |
| mpHNSCC-002 metastatic LN | — | 1300 | 4.418 | 1.541 | 10.588 | 12.205 | 0.856 | 0.336 |
| mpHNSCC-002 metastatic LN | — | 1500 | 4.545 | 1.524 | 10.659 | 13.065 | 0.886 | 0.329 |
| mpHNSCC-002 metastatic LN | — | 1700 | 4.528 | 1.522 | 10.609 | 12.953 | 0.885 | 0.271 |
| mpHNSCC-002 metastatic LN | — | 1900 | 4.755 | 1.488 | 10.578 | 14.515 | 0.941 | 0.277 |
| mpHNSCC-002 metastatic LN | — | 2100 | 4.196 | 2.070 | 12.937 | 9.319 | 0.624 | 0.226 |
| mpHNSCC-002 metastatic LN | — | 2500 | 5.737 | 4.974 | 29.764 | 11.309 | 0.363 | 0.166 |
| mpHNSCC-002 metastatic LN | — | 2900 | 5.023 | 1.689 | 13.063 | 15.943 | 0.884 | 0.144 |
| mpHNSCC-002 metastatic LN | — | 3300 | 5.129 | 1.504 | 11.067 | 17.039 | 0.995 | 0.158 |
| mpHNSCC-009 metastatic LN | — | 900 | 5.022 | 1.286 | 8.055 | 16.313 | 1.112 | 0.894 |
| mpHNSCC-009 metastatic LN | — | 1100 | 5.903 | 1.746 | 14.885 | 22.547 | 0.987 | 0.607 |
| mpHNSCC-009 metastatic LN | — | 1300 | 6.393 | 1.789 | 15.784 | 26.545 | 1.034 | 0.485 |
| mpHNSCC-009 metastatic LN | — | 1500 | 6.331 | 1.815 | 16.205 | 26.007 | 1.014 | 0.390 |
| mpHNSCC-009 metastatic LN | — | 1700 | 5.897 | 1.829 | 16.051 | 22.360 | 0.948 | 0.320 |
| mpHNSCC-009 metastatic LN | — | 1900 | 6.681 | 1.867 | 17.194 | 28.990 | 1.035 | 0.338 |
| mpHNSCC-009 metastatic LN | — | 2100 | 6.331 | 2.023 | 19.342 | 25.630 | 0.924 | 0.274 |
| mpHNSCC-009 metastatic LN | — | 2500 | 6.132 | 1.588 | 12.325 | 24.353 | 1.102 | 0.211 |
| mpHNSCC-009 metastatic LN | — | 2900 | 6.349 | 2.303 | 22.886 | 24.871 | 0.827 | 0.197 |
| mpHNSCC-009 metastatic LN | — | 3300 | 6.835 | 2.334 | 24.680 | 29.389 | 0.872 | 0.225 |
| mpHNSCC-012 metastatic LN | — | 900 | 3.541 | 1.428 | 7.926 | 7.410 | 0.752 | 0.460 |
| mpHNSCC-012 metastatic LN | — | 1100 | 3.487 | 1.288 | 7.046 | 7.452 | 0.813 | 0.370 |
| mpHNSCC-012 metastatic LN | — | 1300 | 3.552 | 1.182 | 6.442 | 7.995 | 0.893 | 0.233 |
| mpHNSCC-012 metastatic LN | — | 1500 | 3.444 | 1.220 | 6.552 | 7.377 | 0.845 | 0.220 |
| mpHNSCC-012 metastatic LN | — | 1700 | 3.530 | 1.252 | 6.893 | 7.750 | 0.844 | 0.203 |
| mpHNSCC-012 metastatic LN | — | 1900 | 3.556 | 1.270 | 7.054 | 7.843 | 0.838 | 0.180 |
| mpHNSCC-012 metastatic LN | — | 2100 | 3.604 | 1.262 | 7.078 | 8.110 | 0.853 | 0.127 |
| mpHNSCC-012 metastatic LN | — | 2500 | 4.140 | 1.274 | 7.818 | 11.036 | 0.954 | 0.096 |
| mpHNSCC-012 metastatic LN | — | 2900 | 3.961 | 1.328 | 8.092 | 9.920 | 0.887 | 0.096 |
| mpHNSCC-012 metastatic LN | — | 3300 | 3.785 | 1.371 | 8.122 | 8.845 | 0.828 | 0.123 |
| mpHNSCC-014 metastatic LN | — | 1300 | 8.982 | 1.330 | 2.698 | 37.334 | 1.499 | 0.426 |
| mpHNSCC-014 metastatic LN | — | 1500 | 8.049 | 1.461 | 4.800 | 36.321 | 1.439 | 0.383 |
| mpHNSCC-014 metastatic LN | — | 1700 | 7.534 | 2.123 | 22.218 | 36.857 | 1.028 | 0.351 |
| mpHNSCC-014 metastatic LN | — | 1900 | 7.995 | 1.856 | 15.679 | 40.563 | 1.202 | 0.341 |
| mpHNSCC-014 metastatic LN | — | 2100 | 6.203 | 1.419 | 9.023 | 24.313 | 1.215 | 0.261 |
| mpHNSCC-014 metastatic LN | — | 2500 | 5.247 | 1.225 | 6.875 | 17.495 | 1.196 | 0.216 |
| mpHNSCC-014 metastatic LN | — | 2900 | 5.160 | 1.294 | 8.075 | 17.172 | 1.131 | 0.212 |
| mpHNSCC-014 metastatic LN | — | 3300 | 4.849 | 1.162 | 6.323 | 15.036 | 1.173 | 0.223 |
| mpHNSCC-016 metastatic LN | — | 900 | 5.003 | 3.327 | 21.109 | 10.716 | 0.470 | 0.462 |
| mpHNSCC-016 metastatic LN | — | 1100 | 6.302 | 2.133 | 20.745 | 25.049 | 0.879 | 0.206 |
| mpHNSCC-016 metastatic LN | — | 1300 | 6.988 | 2.073 | 20.972 | 31.597 | 0.985 | 0.194 |
| mpHNSCC-016 metastatic LN | — | 1500 | 6.890 | 2.495 | 26.908 | 29.311 | 0.828 | 0.300 |
| mpHNSCC-016 metastatic LN | — | 1700 | 6.792 | 2.305 | 24.187 | 29.075 | 0.877 | 0.364 |
| mpHNSCC-016 metastatic LN | — | 1900 | 6.595 | 1.945 | 18.498 | 28.157 | 0.990 | 0.413 |
| mpHNSCC-016 metastatic LN | — | 2100 | 6.491 | 1.594 | 12.115 | 27.082 | 1.150 | 0.273 |
| mpHNSCC-016 metastatic LN | — | 2500 | 6.374 | 1.962 | 18.528 | 26.162 | 0.955 | 0.154 |
| mpHNSCC-016 metastatic LN | — | 2900 | 7.256 | 1.890 | 17.483 | 34.114 | 1.097 | 0.095 |
| mpHNSCC-016 metastatic LN | — | 3300 | 6.862 | 1.749 | 14.864 | 30.442 | 1.117 | 0.106 |
| mpHNSCC-017 metastatic LN | — | 900 | 4.303 | 2.552 | 14.954 | 8.650 | 0.524 | 0.605 |
| mpHNSCC-017 metastatic LN | — | 1100 | 4.351 | 2.513 | 15.110 | 9.009 | 0.538 | 0.585 |
| mpHNSCC-017 metastatic LN | — | 1300 | 4.505 | 2.333 | 15.341 | 10.415 | 0.596 | 0.348 |
| mpHNSCC-017 metastatic LN | — | 1500 | 4.585 | 2.697 | 16.917 | 9.875 | 0.528 | 0.286 |
| mpHNSCC-017 metastatic LN | — | 1700 | 4.676 | 2.597 | 17.138 | 10.703 | 0.558 | 0.260 |
| mpHNSCC-017 metastatic LN | — | 1900 | 4.791 | 2.569 | 17.676 | 11.507 | 0.577 | 0.293 |
| mpHNSCC-017 metastatic LN | — | 2100 | 5.029 | 2.500 | 18.673 | 13.321 | 0.620 | 0.218 |
| mpHNSCC-017 metastatic LN | — | 2500 | 5.042 | 2.273 | 17.597 | 14.192 | 0.679 | 0.099 |
| mpHNSCC-017 metastatic LN | — | 2900 | 5.153 | 2.480 | 19.213 | 14.268 | 0.639 | 0.102 |
| mpHNSCC-017 metastatic LN | — | 3300 | 5.199 | 2.457 | 19.332 | 14.685 | 0.650 | 0.144 |
| mpHNSCC-018 metastatic LN | — | 1500 | 9.677 | 1.818 | 8.949 | 53.765 | 1.406 | 0.414 |
| mpHNSCC-018 metastatic LN | — | 1700 | 9.460 | 2.031 | 16.790 | 55.258 | 1.276 | 0.311 |
| mpHNSCC-018 metastatic LN | — | 1900 | 8.570 | 2.283 | 25.650 | 47.658 | 1.077 | 0.272 |
| mpHNSCC-018 metastatic LN | — | 2100 | 8.774 | 2.461 | 29.871 | 49.993 | 1.032 | 0.238 |
| mpHNSCC-018 metastatic LN | — | 2500 | 8.923 | 2.443 | 29.446 | 51.716 | 1.053 | 0.175 |
| mpHNSCC-018 metastatic LN | — | 2900 | 8.644 | 2.526 | 31.251 | 48.406 | 0.998 | 0.221 |
| mpHNSCC-018 metastatic LN | — | 3300 | 8.177 | 2.385 | 27.876 | 43.328 | 0.999 | 0.349 |
| mpHNSCC-018 metastatic LN | — | 4700 | 7.912 | 2.471 | 29.196 | 40.203 | 0.943 | 0.091 |
| mpHNSCC-018 metastatic LN | — | 5000 | 7.857 | 2.452 | 28.762 | 39.660 | 0.943 | 0.082 |
| mpHNSCC-001 tumor | N+ | 900 | 5.711 | 2.061 | 18.411 | 20.159 | 0.831 | 0.463 |
| mpHNSCC-001 tumor | N+ | 1100 | 5.436 | 2.221 | 18.899 | 17.355 | 0.743 | 0.440 |
| mpHNSCC-001 tumor | N+ | 1300 | 5.409 | 1.596 | 12.443 | 18.936 | 0.989 | 0.418 |
| mpHNSCC-001 tumor | N+ | 1500 | 5.174 | 1.592 | 12.200 | 17.239 | 0.955 | 0.387 |
| mpHNSCC-001 tumor | N+ | 1700 | 5.301 | 1.911 | 15.847 | 17.377 | 0.831 | 0.338 |
| mpHNSCC-001 tumor | N+ | 1900 | 5.214 | 1.620 | 12.580 | 17.476 | 0.947 | 0.373 |
| mpHNSCC-001 tumor | N+ | 2100 | 5.576 | 1.772 | 14.894 | 19.901 | 0.928 | 0.297 |
| mpHNSCC-001 tumor | N+ | 2500 | 5.481 | 1.444 | 10.232 | 19.483 | 1.087 | 0.261 |
| mpHNSCC-001 tumor | N+ | 2900 | 5.792 | 2.190 | 19.921 | 20.380 | 0.797 | 0.278 |
| mpHNSCC-001 tumor | N+ | 3300 | 5.641 | 1.325 | 8.088 | 20.257 | 1.191 | 0.267 |
| mpHNSCC-002 tumor | N+ | 900 | 3.490 | 1.272 | 6.951 | 7.505 | 0.824 | 0.359 |
| mpHNSCC-002 tumor | N+ | 1100 | 3.499 | 1.146 | 6.117 | 7.785 | 0.905 | 0.305 |
| mpHNSCC-002 tumor | N+ | 1300 | 3.477 | 1.069 | 5.508 | 7.785 | 0.955 | 0.226 |
| mpHNSCC-002 tumor | N+ | 1500 | 3.463 | 1.064 | 5.454 | 7.724 | 0.956 | 0.232 |
| mpHNSCC-002 tumor | N+ | 1700 | 3.466 | 1.051 | 5.349 | 7.751 | 0.967 | 0.204 |
| mpHNSCC-002 tumor | N+ | 1900 | 3.754 | 1.001 | 4.931 | 9.143 | 1.076 | 0.212 |
| mpHNSCC-002 tumor | N+ | 2100 | 4.085 | 0.977 | 4.473 | 10.667 | 1.174 | 0.204 |
| mpHNSCC-002 tumor | N+ | 2500 | 3.500 | 1.011 | 5.018 | 7.942 | 1.007 | 0.116 |
| mpHNSCC-002 tumor | N+ | 2900 | 3.931 | 1.153 | 6.509 | 10.010 | 0.994 | 0.111 |
| mpHNSCC-002 tumor | N+ | 3300 | 3.941 | 1.072 | 5.673 | 10.086 | 1.058 | 0.126 |
| mpHNSCC-003 tumor | N- | 900 | 6.830 | 3.510 | 35.120 | 24.058 | 0.601 | 0.310 |
| mpHNSCC-003 tumor | N- | 1100 | 6.669 | 3.362 | 33.114 | 23.219 | 0.612 | 0.350 |
| mpHNSCC-003 tumor | N- | 1300 | 6.910 | 3.079 | 32.762 | 26.823 | 0.686 | 0.281 |
| mpHNSCC-003 tumor | N- | 1500 | 6.803 | 3.382 | 34.176 | 24.381 | 0.620 | 0.263 |
| mpHNSCC-003 tumor | N- | 1700 | 6.895 | 3.357 | 34.661 | 25.369 | 0.632 | 0.243 |
| mpHNSCC-003 tumor | N- | 1900 | 7.016 | 3.133 | 33.836 | 27.625 | 0.685 | 0.272 |
| mpHNSCC-003 tumor | N- | 2100 | 7.140 | 3.067 | 34.007 | 29.212 | 0.710 | 0.172 |
| mpHNSCC-003 tumor | N- | 2500 | 7.978 | 2.763 | 34.221 | 39.882 | 0.862 | 0.110 |
| mpHNSCC-003 tumor | N- | 2900 | 7.522 | 3.162 | 37.080 | 32.767 | 0.724 | 0.101 |
| mpHNSCC-003 tumor | N- | 3300 | 7.568 | 2.817 | 33.458 | 35.006 | 0.808 | 0.123 |
| mpHNSCC-004 tumor | N- | 900 | 4.374 | 1.259 | 7.795 | 12.410 | 1.010 | 0.560 |
| mpHNSCC-004 tumor | N- | 1100 | 4.451 | 1.319 | 8.490 | 12.819 | 0.986 | 0.442 |
| mpHNSCC-004 tumor | N- | 1300 | 4.690 | 1.290 | 8.208 | 14.288 | 1.049 | 0.355 |
| mpHNSCC-004 tumor | N- | 1500 | 4.915 | 1.452 | 10.302 | 15.636 | 0.988 | 0.278 |
| mpHNSCC-004 tumor | N- | 1700 | 5.171 | 1.309 | 8.306 | 17.269 | 1.123 | 0.264 |
| mpHNSCC-004 tumor | N- | 1900 | 5.449 | 1.454 | 10.408 | 19.267 | 1.075 | 0.257 |
| mpHNSCC-004 tumor | N- | 2100 | 5.373 | 1.398 | 9.573 | 18.703 | 1.098 | 0.177 |
| mpHNSCC-004 tumor | N- | 2500 | 5.978 | 1.494 | 10.747 | 23.036 | 1.134 | 0.106 |
| mpHNSCC-004 tumor | N- | 2900 | 8.810 | 1.341 | 1.656 | 37.034 | 1.526 | 0.117 |
| mpHNSCC-004 tumor | N- | 3300 | 6.430 | 1.475 | 9.762 | 26.145 | 1.213 | 0.124 |
| mpHNSCC-005 tumor | N- | 900 | 3.366 | 1.409 | 7.398 | 6.577 | 0.727 | 0.452 |
| mpHNSCC-005 tumor | N- | 1100 | 3.501 | 1.535 | 8.311 | 6.947 | 0.696 | 0.340 |
| mpHNSCC-005 tumor | N- | 1300 | 3.604 | 1.535 | 8.602 | 7.473 | 0.715 | 0.256 |
| mpHNSCC-005 tumor | N- | 1500 | 3.740 | 1.542 | 9.019 | 8.180 | 0.737 | 0.237 |
| mpHNSCC-005 tumor | N- | 1700 | 3.832 | 1.586 | 9.496 | 8.573 | 0.734 | 0.229 |
| mpHNSCC-005 tumor | N- | 1900 | 3.936 | 1.506 | 9.311 | 9.368 | 0.788 | 0.246 |
| mpHNSCC-005 tumor | N- | 2100 | 4.001 | 1.518 | 9.536 | 9.713 | 0.795 | 0.179 |
| mpHNSCC-005 tumor | N- | 2500 | 4.225 | 1.607 | 10.664 | 10.821 | 0.793 | 0.110 |
| mpHNSCC-005 tumor | N- | 2900 | 4.189 | 1.470 | 9.587 | 10.952 | 0.852 | 0.111 |
| mpHNSCC-005 tumor | N- | 3300 | 4.456 | 1.518 | 10.458 | 12.500 | 0.874 | 0.132 |
| mpHNSCC-006 tumor | N- | 900 | 5.636 | 1.659 | 13.454 | 20.567 | 0.991 | 0.615 |
| mpHNSCC-006 tumor | N- | 1100 | 5.209 | 1.652 | 12.959 | 17.379 | 0.930 | 0.547 |
| mpHNSCC-006 tumor | N- | 1300 | 5.120 | 1.744 | 13.802 | 16.501 | 0.874 | 0.407 |
| mpHNSCC-006 tumor | N- | 1500 | 4.970 | 1.694 | 13.017 | 15.547 | 0.874 | 0.342 |
| mpHNSCC-006 tumor | N- | 1700 | 4.768 | 1.669 | 12.386 | 14.193 | 0.853 | 0.288 |
| mpHNSCC-006 tumor | N- | 1900 | 4.629 | 1.530 | 10.849 | 13.596 | 0.897 | 0.312 |
| mpHNSCC-006 tumor | N- | 2100 | 4.625 | 1.674 | 12.114 | 13.212 | 0.829 | 0.189 |
| mpHNSCC-006 tumor | N- | 2500 | 5.027 | 1.591 | 12.030 | 16.193 | 0.932 | 0.130 |
| mpHNSCC-006 tumor | N- | 2900 | 4.847 | 1.628 | 12.151 | 14.848 | 0.885 | 0.098 |
| mpHNSCC-006 tumor | N- | 3300 | 4.969 | 1.695 | 13.026 | 15.542 | 0.873 | 0.107 |
| mpHNSCC-011 tumor | N- | 900 | 5.601 | 1.823 | 15.547 | 19.984 | 0.910 | 0.402 |
| mpHNSCC-011 tumor | N- | 1100 | 5.293 | 1.849 | 15.223 | 17.507 | 0.855 | 0.330 |
| mpHNSCC-011 tumor | N- | 1300 | 5.250 | 1.830 | 14.940 | 17.230 | 0.856 | 0.255 |
| mpHNSCC-011 tumor | N- | 1500 | 5.325 | 1.804 | 14.831 | 17.882 | 0.878 | 0.245 |
| mpHNSCC-011 tumor | N- | 1700 | 5.469 | 1.660 | 13.339 | 19.295 | 0.966 | 0.223 |
| mpHNSCC-011 tumor | N- | 1900 | 5.465 | 1.837 | 15.465 | 18.875 | 0.884 | 0.246 |
| mpHNSCC-011 tumor | N- | 2100 | 5.411 | 1.689 | 13.654 | 18.809 | 0.943 | 0.139 |
| mpHNSCC-011 tumor | N- | 2500 | 5.576 | 1.502 | 11.113 | 20.187 | 1.068 | 0.090 |
| mpHNSCC-011 tumor | N- | 2900 | 5.875 | 1.622 | 12.983 | 22.416 | 1.046 | 0.087 |
| mpHNSCC-011 tumor | N- | 3300 | 6.057 | 1.944 | 17.820 | 23.437 | 0.921 | 0.109 |
| mpHNSCC-013 tumor | N- | 1900 | 11.943 | 4.167 | 77.401 | 89.164 | 0.856 | 0.539 |
| mpHNSCC-013 tumor | N- | 2100 | 12.177 | 4.666 | 89.256 | 89.600 | 0.787 | 0.337 |
| mpHNSCC-013 tumor | N- | 2500 | 12.066 | 5.205 | 97.445 | 83.225 | 0.707 | 0.159 |
| mpHNSCC-013 tumor | N- | 2900 | 11.456 | 4.992 | 88.531 | 74.632 | 0.700 | 0.149 |
| mpHNSCC-013 tumor | N- | 3300 | 11.247 | 4.321 | 76.339 | 76.354 | 0.785 | 0.200 |
| mpHNSCC-014 tumor | N+ | 900 | 3.025 | 1.921 | 7.595 | 4.062 | 0.491 | 0.451 |
| mpHNSCC-014 tumor | N+ | 1100 | 3.084 | 2.049 | 8.016 | 4.075 | 0.470 | 0.715 |
| mpHNSCC-014 tumor | N+ | 1300 | 3.279 | 1.979 | 8.745 | 4.959 | 0.516 | 0.559 |
| mpHNSCC-014 tumor | N+ | 1500 | 3.455 | 2.010 | 9.560 | 5.652 | 0.534 | 0.475 |
| mpHNSCC-014 tumor | N+ | 1700 | 3.544 | 2.163 | 10.262 | 5.744 | 0.510 | 0.375 |
| mpHNSCC-014 tumor | N+ | 1900 | 3.569 | 2.179 | 10.410 | 5.824 | 0.510 | 0.355 |
| mpHNSCC-014 tumor | N+ | 2100 | 3.712 | 2.220 | 11.166 | 6.395 | 0.520 | 0.223 |
| mpHNSCC-014 tumor | N+ | 2500 | 3.797 | 2.313 | 11.770 | 6.602 | 0.511 | 0.166 |
| mpHNSCC-014 tumor | N+ | 2900 | 4.193 | 2.275 | 13.623 | 8.742 | 0.571 | 0.170 |
| mpHNSCC-014 tumor | N+ | 3300 | 4.285 | 2.411 | 14.481 | 8.906 | 0.551 | 0.201 |
| mpHNSCC-015 tumor | N- | 900 | 5.333 | 3.349 | 23.501 | 12.731 | 0.496 | 0.225 |
| mpHNSCC-015 tumor | N- | 1100 | 5.643 | 3.563 | 26.366 | 14.194 | 0.494 | 0.266 |
| mpHNSCC-015 tumor | N- | 1300 | 5.904 | 3.435 | 27.918 | 16.511 | 0.534 | 0.261 |
| mpHNSCC-015 tumor | N- | 1500 | 6.049 | 3.438 | 28.996 | 17.625 | 0.546 | 0.361 |
| mpHNSCC-015 tumor | N- | 1700 | 6.281 | 3.431 | 30.674 | 19.531 | 0.567 | 0.328 |
| mpHNSCC-015 tumor | N- | 1900 | 6.676 | 3.313 | 32.879 | 23.507 | 0.621 | 0.319 |
| mpHNSCC-015 tumor | N- | 2100 | 6.790 | 3.481 | 34.657 | 23.814 | 0.602 | 0.199 |
| mpHNSCC-015 tumor | N- | 2500 | 7.393 | 3.743 | 40.796 | 28.461 | 0.609 | 0.141 |
| mpHNSCC-015 tumor | N- | 2900 | 7.351 | 3.741 | 40.449 | 28.046 | 0.606 | 0.182 |
| mpHNSCC-015 tumor | N- | 3300 | 7.696 | 4.174 | 45.884 | 29.465 | 0.571 | 0.190 |
| mpHNSCC-016 tumor | N+ | 900 | 2.068 | 0.627 | 1.903 | 2.759 | 0.967 | 0.430 |
| mpHNSCC-016 tumor | N+ | 1100 | 2.277 | 0.595 | 1.735 | 3.360 | 1.094 | 0.225 |
| mpHNSCC-016 tumor | N+ | 1300 | 2.499 | 0.585 | 1.572 | 3.973 | 1.194 | 0.139 |
| mpHNSCC-016 tumor | N+ | 1500 | 2.545 | 0.559 | 1.322 | 4.034 | 1.254 | 0.124 |
| mpHNSCC-016 tumor | N+ | 1700 | 2.427 | 0.969 | 3.688 | 3.498 | 0.759 | 0.140 |
| mpHNSCC-016 tumor | N+ | 1900 | 2.827 | 0.692 | 2.276 | 5.132 | 1.153 | 0.101 |
| mpHNSCC-016 tumor | N+ | 2100 | 2.844 | 0.686 | 2.215 | 5.182 | 1.167 | 0.095 |
| mpHNSCC-016 tumor | N+ | 2500 | 3.014 | 1.025 | 4.771 | 5.721 | 0.876 | 0.066 |
| mpHNSCC-016 tumor | N+ | 2900 | 13.470 | 3.932 | 75.757 | 117.561 | 0.998 | 0.097 |
| mpHNSCC-017 tumor | N+ | 900 | 3.571 | 1.597 | 8.772 | 7.151 | 0.684 | 0.504 |
| mpHNSCC-017 tumor | N+ | 1100 | 3.716 | 1.627 | 9.351 | 7.831 | 0.697 | 0.328 |
| mpHNSCC-017 tumor | N+ | 1300 | 3.754 | 1.722 | 9.878 | 7.791 | 0.668 | 0.253 |
| mpHNSCC-017 tumor | N+ | 1500 | 3.912 | 1.750 | 10.530 | 8.578 | 0.684 | 0.265 |
| mpHNSCC-017 tumor | N+ | 1700 | 4.178 | 1.650 | 10.819 | 10.415 | 0.766 | 0.234 |
| mpHNSCC-017 tumor | N+ | 1900 | 4.089 | 1.802 | 11.382 | 9.452 | 0.693 | 0.246 |
| mpHNSCC-017 tumor | N+ | 2100 | 4.362 | 1.692 | 11.591 | 11.441 | 0.779 | 0.142 |
| mpHNSCC-017 tumor | N+ | 2500 | 4.569 | 1.616 | 11.515 | 12.996 | 0.846 | 0.062 |
| mpHNSCC-017 tumor | N+ | 2900 | 4.894 | 1.642 | 12.364 | 15.140 | 0.886 | 0.082 |
| mpHNSCC-017 tumor | N+ | 3300 | 5.037 | 1.663 | 12.828 | 16.101 | 0.898 | 0.073 |
| mpHNSCC-018 tumor | N+ | 900 | 2.216 | 0.747 | 2.551 | 3.100 | 0.882 | 0.376 |
| mpHNSCC-018 tumor | N+ | 1100 | 2.398 | 0.724 | 2.546 | 3.714 | 0.970 | 0.269 |
| mpHNSCC-018 tumor | N+ | 1300 | 2.490 | 0.713 | 2.503 | 4.024 | 1.014 | 0.191 |
| mpHNSCC-018 tumor | N+ | 1500 | 2.665 | 0.688 | 2.310 | 4.599 | 1.105 | 0.205 |
| mpHNSCC-018 tumor | N+ | 1700 | 2.645 | 0.737 | 2.678 | 4.545 | 1.038 | 0.166 |
| mpHNSCC-018 tumor | N+ | 1900 | 2.653 | 0.758 | 2.826 | 4.567 | 1.017 | 0.151 |
| mpHNSCC-018 tumor | N+ | 2100 | 2.824 | 0.772 | 2.940 | 5.179 | 1.054 | 0.107 |
| mpHNSCC-018 tumor | N+ | 2500 | 2.928 | 0.823 | 3.343 | 5.567 | 1.030 | 0.057 |
| mpHNSCC-018 tumor | N+ | 2900 | 2.901 | 0.739 | 2.653 | 5.441 | 1.117 | 0.078 |
| mpHNSCC-018 tumor | N+ | 3300 | 3.506 | 1.013 | 5.039 | 7.972 | 1.007 | 0.087 |

**Supplementary Table S4 — Nomenclature and symbol conventions**

*Symbols used consistently across the main manuscript, all figures and tables, the data workbook and this Supplement.*

| **Symbol** | **Quantity** | **Unit** | **Frequency-resolved** | **Note** |
| --- | --- | --- | --- | --- |
| SWS | shear-wave speed | m/s | yes | used for both ttMRE and ultrasound |
| PR | penetration rate | m/s | yes | reciprocal spatial attenuation as a velocity |
| G′ | storage modulus | kPa | yes | real part of G* |
| G″ | loss modulus | kPa | yes | imaginary part of G* |
| φ | loss angle | rad | yes | arctan(G″/G′) |
| σ | Bessel-fit residual | dimensionless | yes | goodness of fit of the global Bessel fit |
| μ | shear modulus (stiffness) | kPa | no | spring-pot fit, pixel-wise |
| α | power-law exponent | dimensionless | no | spring-pot fit, pixel-wise; range 0–1 |
| H | Shannon entropy | nats | no | natural logarithm, 30 bins |
| ttMRE | tabletop MR elastography | — | — | ex vivo, 0.5 T benchtop system |
| USE | ultrasound shear-wave elastography | — | — | ARFI-based, in vivo and ex vivo |
| N− / N+ | node-negative / node-positive primary tumor | — | — |  |
| OrientationJ | ImageJ plugin for fiber-orientation overlays | — | — | illustrative only; no orientation statistics derived |
